# Discovery of genomic and transcriptomic pleiotropy between kidney function and soluble receptor for advanced glycation end-products using correlated meta-analyses: The Long Life Family Study (LLFS)

**DOI:** 10.1101/2023.12.27.23300583

**Authors:** Mary F. Feitosa, Shiow J. Lin, Sandeep Acharya, Bharat Thyagarajan, Mary K. Wojczynski, Allison L. Kuipers, Alexander Kulminski, Kaare Christensen, Joseph M. Zmuda, Michael R. Brent, Michael A. Province

## Abstract

Patients with chronic kidney disease (CKD) have increased oxidative stress and chronic inflammation, which may escalate the production of advanced glycation end-products (AGE). High soluble receptor for AGE (sRAGE) and low estimated glomerular filtration rate (eGFR) levels are associated with CKD and aging. We evaluated whether eGFR calculated from creatinine and cystatin C share pleiotropic genetic factors with sRAGE. We employed whole-genome sequencing and correlated meta-analyses on combined genomewide association study (GWAS) *p*-values in 4,182 individuals (age range: 24-110) from the Long Life Family Study (LLFS). We also conducted transcriptome-wide association studies (TWAS) on whole blood in a subset of 1,209 individuals. We identified 59 pleiotropic GWAS loci (*p*<5×10^-8^) and 17 TWAS genes (Bonferroni-*p*<2.73×10^-6^) for eGFR traits and sRAGE. TWAS genes, *LSP1* and *MIR23AHG*, were associated with eGFR and sRAGE located within GWAS loci, lncRNA-*KCNQ1OT1* and *CACNA1A/CCDC130*, respectively. GWAS variants were eQTLs in the kidney glomeruli and tubules, and GWAS genes predicted kidney carcinoma. TWAS genes harbored eQTLs in the kidney, predicted kidney carcinoma, and connected enhancer-promoter variants with kidney function-related phenotypes at *p*<5×10^-8^. Additionally, higher allele frequencies of protective variants for eGFR traits were detected in LLFS than in ALFA-Europeans and TOPMed, suggesting better kidney function in healthy-aging LLFS than in general populations. Integrating genomic annotation and transcriptional gene activity revealed the enrichment of genetic elements in kidney function and kidney diseases. The identified pleiotropic loci and gene expressions for eGFR and sRAGE suggest their underlying shared genetic effects and highlight their roles in kidney- and aging-related signaling pathways.

## Introduction

Patients with chronic kidney disease (CKD) have increased chronic inflammation and oxidative stress that may escalate the production of advanced glycation end-products (AGEs). Increased AGEs and decreased kidney clearance may lead to the accumulation of AGEs and the imbalance between oxidant/antioxidant capacities.^1, 2^ AGEs and the receptor for AGE (RAGE) are implicated in CKD progression and CKD-related complications; however, the precise mechanisms of action of their relation with kidney function are not fully understood. Serum levels of soluble RAGE (sRAGE) are a decoy receptor that suppresses membrane-bound RAGE activation and AGE-RAGE-related toxicity and has been proposed as a therapeutic agent targeting vascular inflammation to prevent cardiovascular disease.^3^ In contrast, the direction of the association of sRAGE and kidney disease seems to be the inverse of other chronic diseases, in which high levels of sRAGE are associated with worse kidney function.^2, 3^ High circulating sRAGE levels were associated with incident CKD and end-stage renal disease (ESRD) risk^4^ and inversely associated with the estimated glomerular filtration rate (eGFR).^4, 5^ However, the association of CKD and ESRD with sRAGE was not significant after adjusting for baseline eGFR in the ARIC study. Whether the sRAGE levels are directly affected by the decline in eGFR or if high sRAGE levels directly impact kidney function remains undetermined^4^.

In clinical practice, the eGFR calculated from serum creatinine (eGFRcr) has been the marker of choice for kidney function. Serum creatinine is known to be influenced by factors other than GFR, such as muscle mass, diet, activity, and age.^6–11^ The eGFR calculated from serum cystatin C level (eGFRcys) is less dependent on muscle mass, but it may be influenced by inflammation, obesity, and diabetes.^8, 9, 11^ Several studies have reported that eGFRcr and eGFRcys are often divergent, particularly in older adults.^6–10^ Serum cystatin C level strengthens the association between the eGFR and the risks of ESRD and death across diverse populations. It is demonstrated as a biomarker of aging because its high levels are associated with worse physical disabilities and comorbidities among older adults.^7^ Lower eGFRcys levels are also associated with a higher risk for frailty, hospitalization rates, and mortality, while lower eGFRcr levels are not. Thus, eGFRcys may complement eGFRcr for managing kidney function. The Kidney Disease Improving Global Outcomes (KDIGO) also recommended using the eGFRcys for confirmatory testing of eGFRcr.^12^

Both genetic and environmental factors contribute to kidney function. In twin studies, the broad-sense heritability estimates were 54% for eGFRcr and 60% for eGFRcys.^13^ About 420 loci have been identified from genomewide association studies (GWAS) for eGFRcr.^14, 15^ The recent CKD Genetics Consortium and UK Biobank study reported the eGFRcr variance of 9.8% by 634 independent signals,^14^ which suggests that the explained variance is still missing heritability. Low frequency and rare variants from whole genome sequencing (WGS) data may help to recover the eGFR heritability^16^.

The transcriptome-wide association study (TWAS) is a gene-based association approach that detects the association between transcript levels of a gene and a trait. TWAS may help to prioritize GWAS signals in which associated genetic variants may regulate gene expression levels, modulating the disease risk or modifying the trait levels. On whole blood from the GTEx and from micro-dissected kidney tubules, TWAS of eGFRcr and eGFRcys identified 849 and 416 transcript associations, respectively, and 229 transcript associations were found across eGFRcr and eGFRcys. Colocalizing expression quantitative trait loci (eQTL) and GWAS for kidney function resulted in 214 unique genes.^17^

The discovery of genomic and transcriptomic pleiotropy between kidney function and sRAGE can provide etiological insights into systemic inflammation and CKD, which may increase the risk of developing kidney diabetes, ESRD, and mortality. The correlated meta-analysis (CMA)^18, 19^ can enhance the ability to detect pleiotropic genetic effects on correlated phenotypes by empirically estimating the covariance among GWAS or TWAS. The CMA approach corrects for the false-positive signals arising from non-independent data in the combined GWAS or TWAS *p*-values.

In the current study, we had three main goals. Firstly, we aimed to identify pleiotropic WGS variants among GWAS of eGFRcr, eGFRcys, and sRAGE through CMA in the Long Life Family Study, which recruited participants in the upper generation with exceptional family longevity. Secondly, we wanted to identify the association between kidney function traits with expression protein-coding genes and long-intergenic non-protein coding RNA (lncRNA) employing TWAS for eGFRcr, eGFRcys, and sRAGE by assessing RNA-sequencing (RNA-seq) on whole blood. We then searched for pleiotropic genes among these traits through CMA. Lastly, we conducted bioinformatic analyses and reviewed the literature to determine whether the identified variants and genes may tag potentially functional genome elements that can be implicated in kidney and aging biological pathways.

## Material and methods

### Study Data

The Long Life Family Study (LLFS) is a longitudinal, population-based multigenerational family study designed to investigate genetic, behavioral, and environmental factors in families exhibiting exceptional longevity. Families were sampled from four clinical centers: Boston University Medical Center in Boston, MA; Columbia College of Physicians and Surgeons in New York City, NY; the University of Pittsburgh in Pittsburgh, PA, USA; and the University of Southern Denmark, Odense, Denmark. The characteristics, recruitment, eligibility, and enrollment were previously described.^20, 21^ The first clinical exam started in 2006 and recruited 4,953 individuals in 539 two-generation families clustered for exceptional survival in the upper generation. In the second clinical exam (2014-2017), 2,933 individuals from 528 families were revisited. The third clinical exam (2021-) is still recruiting individuals from the second exam and new ones from the grandchildren generation. All participants signed informed consent. The Institutional Review Boards approved all study procedures of participating institutions.

### Traits

Serum creatinine was measured in EDTA plasma using the enzymatic method on a Roche Modular P Chemistry Analyzer (Roche Diagnostics Corporation). The procedure was calibrated using the National Institute of Standards and Technology guide to reference material SRM 909b (Isotope Dilution Mass Spectroscopy). The laboratory inter-assay coefficient of variation (CV) was 2.3%. Cystatin C was measured in serum using Gentian Cystatin C reagent (Gentian AS, Moss, Norway) on the Roche Cobas 6000 Chemistry analyzer (Roche Diagnostics Corporation). The laboratory inter-assay CVs were 5.6% at 0.76 mg/L and 3.8% at 3.22 mg/L. The lower limit of detection was 0.3 mg/L. The new race-free serum creatinine equation for eGFRcr and serum cystatin C equation for eGFRcys were calculated using the CKD-EPI.^12^ CKD is defined as below 60 ml/min/1.73 m^2^ for both eGFRcr and eGFRcys.

sRAGE was measured in serum using the quantitative sandwich enzyme immunoassay technique of the Human sRAGE ELISA from BioVendor (Asheville, NC). The intensity of the color was measured on a SpectraMax plate reader (Molecular Devices, Sunnyvale, California). The laboratory inter-assay CVs were 13.1% and 6.4% at mean concentrations of 166.3 and 1203 pg/mL for lyophilized manufacturer’s controls and 16.0% at a mean concentration of 359.6 pg/mL for an in-house pooled serum control. The lower limit of detection was 19.2 pg/mL. Levels of sRAGE, eGFRcr, and eGFRcys were log-transformed using a natural logarithm.

Cardiovascular risk factors associated with CKD were defined for coronary heart disease (CHD: self-report of a coronary bypass, myocardial infarction, coronary angioplasty, balloon angioplasty, atherectomy, stent, percutaneous transluminal coronary angioplasty, or percutaneous coronary intervention), type 2 diabetes (T2D: fasting glucose ≥126mg/dL, HbA1c ≥6.5%, taking diabetes medications, self-report of T2D, or having a doctor diagnosis of T2D), and hypertension (systolic blood pressure (BP) >140 mm Hg or diastolic BP>90mmHg or taking BP lowering medications).

### Whole genome sequencing

The WGS in 4,713 LLFS participants was conducted by the McDonnell Genome Institute at Washington University via 150bp reads by Illumina Sequencers. In brief, the sequence alignment to the NCBI-Genome Reference Consortium Human 38 (GRCh38) was implemented by Burrow-Wheeler Aligner. The analytical procedures used for the WGS quality control included Picard to detect duplicates, Genome Analysis Toolkit (GATK) to base quality score recalibration, and SAMtools to compress sequence data using lossless conversion to CRAM format. Additional quality control was performed at the Division of Statistical Genomics at Washington University. GATK HaplotypeCaller was used to call variants from the CRAM files and create individual-level GVCF files. The GATK CombineGVCFs and GenotypeGVCFs tools were used to merge GVCFs and join genotyping, respectively. Poor quality, contaminated, or redundant samples were removed as measured by the FREEMIX statistic (>0.03) and insufficient haploid coverage (<20x). For each sequence call at each variant site, the individual call was filtered out for depth <20 or >300. In addition, the sample discrepancies and duplications were identified and removed by comparing WGS with each individual’s GWAS chip data. Participants with active leukemia or other blood cancer were removed. Family relationships were verified using KING (Kinship-based Inference for Gwas), and samples with Mendelian errors were removed. After quality control, 4,494 participants remained with 57,758,794 WGS autosomal diallelic variants (SNPs and INDELs).

### Genomewide Association Study

To investigate whether genetic variants have effects on kidney function and sRAGE, we conducted GWAS using a mixed linear regression model with an additive effect of the genetic variants. The model accounted for dependency among family members through a pedigree-based kinship matrix as a random effect, which was implemented at the Division of Statistical Genomics pipeline (SAS^®^9.4). Age, age^2^, sex, field centers, and first principal component entered as covariates in the linear regression model for log(eGFRcr), log(eGFRcys), and log(sRAGE). Variants with a low minor allele count (MAC<20), insertions, deletions, and structural variants were excluded. We calculated the genomic control inflation factor (λ) for each GWAS, which included 4,182 individuals of European ancestry with information on age, sex, kidney function, sRAGE, and WGS recruited from the first clinical exam.

### RNA Sequencing and Transcriptome-Wide Association Study

The RNA extraction and transcriptomics were processed at the McDonnell Genome Institute at Washington University. Total RNA was extracted from PAXgene™ Blood RNA tubes using the Qiagen PreAnalytiX PAXgene Blood miRNA Kit (Qiagen, Valencia, CA). The Qiagen QIAcube extraction robot performed the extraction according to the company’s standard operating procedure.

The whole blood deep paired-end RNA sequencing (RNA-seq) design, quality control, oversight, pipelines, and filters were done at the Division of Computational & Data Sciences at Washington University. In brief, a series of bioinformatics pipelines (https://nf-co.re/rnaseq) were executed to align Illumina reads to the human genome sequence GRCh38 with GENCODE annotations and data quality control. Genes with fewer than four counts per million in at least 98.5% of samples and with high intergenic coverage were excluded.

We first estimated the effects of age, sex, field centers, three first gene expression principal components, percentage of intergenic reads, batch effect, white blood cells, platelets, red blood cells, neutrophils, and monocytes on expression levels using a linear model. Then, we performed TWAS on expression residuals by using Mixed Model Analysis for Pedigrees and Populations (MMAP, https://mmap.github.io/), which corrects for familial relationships. To remove inflation and bias often observed in TWAS, we used the Bioconductor Bacon package, which constructs an empirical null distribution using a Gibbs Sampling algorithm by fitting a three-component normal mixture on z-scores.^22^ The Bonferroni-corrected significance threshold (*p*=0.05/18,304 genes) is *p*<2.73×10^-6^. The TWAS analyses used a sample of 1,209 individuals available for the current study.

### Correlated Meta-Analysis

We employed the CMA approach to test whether pleiotropic genetic variants and genes were shared between eGFRcr, eGFRcys, and sRAGE. Details on CMA were previously described.^18, 19^ In brief, CMA empirically estimates the covariance among GWAS (or TWAS) and corrects for the signal inference in the combined GWAS (or TWAS) *p*-value. CMA prevents type 1 error by accounting for all sources of dependencies between multiple genome (or transcriptome) scans under the null, including overlapping individuals, cryptic relatedness, and population structure. The criteria for considering a pleiotropic genetic variant were if the individual GWAS *p*<0.01, CMA GWAS *p*<5×10^-8^, and CMA GWAS *p*<GWAS. A novel GWAS locus was defined if the lead genetic variant (*i.e.*, the most significant SNP at *p*<5×10^-8^) was >500 Kb apart from any lead variant reported in the NHGRI-EBI GWAS catalog (https://www.ebi.ac.uk/gwas/). We considered a pleiotropic gene if the CMA TWAS *p* <2.73×10^-6^ and CMA TWAS *p*<TWAS.

### Bioinformatic Analyses

We selected all variants within 1Mb and in high linkage disequilibrium (LD, variant correlation (r^2^)≥0.8) with CMA GWAS lead variants to examine whether the variants might be tagging regulatory variants. The potentially functional implications of regulatory CMA GWAS variants were accessed using the ENCODE Consortium (https://www.encodeproject.org/) and the Roadmap Epigenome Mapping Consortium (http://www.roadmapepigenomics.org/) initiatives via HaploReg (V4.1, https://pubs.broadinstitute.org/mammals/haploreg/haploreg.php). We interrogated publicly available GWAS studies for kidney function-related phenotypes, sRAGE, and longevity via the NHGRI-EBI GWAS catalog (https://www.ebi.ac.uk/gwas, accessed on 09/20/2023). In addition, relevant biological insights for genes residing within a 1Mb interval of CMA GWAS lead variants were sourced from NCBI (https://www.ncbi.nlm.nih.gov/) and GeneCards (https://www.genecards.org/). To integrate CMA GWAS variants with published eQTL, we searched across multiple tissues from the Genotype-Tissue Expression (GTEx Portal-v8, https://gtexportal.org/home/). We assessed the Human Kidney eQTL Atlas (https://susztaklab.com/Kidney_eQTL/)^23^ for the kidney cell fraction eQTL in glomerular and tubule compartments and kidney eQTLs for variants identified in the meta-analysis. In addition, to connect locus genes to kidney carcinoma, we assessed The Cancer Genome Atlas (TCGA) for kidney renal clear cell carcinoma (KIRC), kidney renal papillary cell carcinoma (KIRP), and kidney chromophobe (KICH) datasets via the NIH National Cancer Institute – Genomic Data Commons Data Portal (https://portal.gdc.cancer.gov/).

We accessed the TCGA database to verify if the CMA TWAS genes predicted KIRC, KIRP, and KICH. To link CMA TWAS genes with regulatory elements that may affect kidney function-related phenotypes, we examined the Human Kidney eQTL Atlas, GeneHancer, and EPRI (enhancer-promoter RNA interaction) maps. GeneHancer incorporates a database genomewide integration of pan-tissue enhancers and promoters and their inferred target genes using the GeneCards framework.^24^ GeneHancer connects enhancers to genes, using tissue co-expression correlation between genes and enhancer RNAs and enhancer-targeted transcription factor genes; eQTLs for variants within enhancers; and a promoter-specific genome conformation assay. The GWAS catalog was also examined for CMA TWAS genes with kidney function-related phenotypes using the predicted gene targets from GeneHancer (GeneExon and/or GWAS) through GeneCards. In addition, we searched for regulatory elements accessing the EPRI maps constructed for H1 human embryonic stem (ES) cells, HeLa, HepG2, K562, IMR90, GM12878, and human neural progenitor cells (NPCs).^25^ For each cell line, chromatin immunoprecipitation followed by sequencing (ChIP–seq) data for histone modifications were used to define enhancer and promoter regions accurately. Genetic variants from the International Cancer Genome Consortium database (ICGC; https://dcc.icgc.org/) and GWAS catalog were accessed to investigate the pathological relevance of the identified EPRIs, constructed from RNA in situ conformation sequencing (RIC-seq) datasets that can accurately assign which enhancers regulate which promoters. The RIC-seq and HiChIP allowed detection enhancer-promoter loops at longer distances. Genotype information and expression data for 53 tissues from the GTEx were also used to examine the regulatory function of EPRIs.

## Results

### Descriptive Analysis

The characteristics of 4,182 participants for kidney function, sRAGE, covariates, and cardiovascular risk factors associated with CKD are presented in Supplementary Table 1. The mean age was 70.4±15.7 years old (range 24 to 110 years), and 55% were women. The mean serum level of sRAGE was 601.8±477.3 pg. The medians of eGFRcr and eGFRcys were 70.6 ml/min/1.73 m^2^ and 78.5 ml/min/1.73 m^2^, respectively. The estimated prevalences of CKD were 28.3% and 30.8% based on equations of eGFRcr and eGFRcys, respectively. The correlation estimate between eGFRcr and GFRcys after the adjustment for covariates was 0.70 (*p*=1.0×10^-60^). eGFRcr and eGFRcys were negatively correlated with sRAGE (r=-0.25, *p*=1.4×10^-37^ and r=-0.30, *p*=2.7×10^-53^, respectively), which suggests genetic sharing effects on the correlated traits.

### Genomewide association study and correlated meta-analysis

We first conducted GWAS analysis for eGFRcr, eGFRcys, and sRAGE. Supplementary Figure 1 depicts each trait’s observed versus expected GWAS −log10 (*p*-value) distribution. The estimated genomic inflation factors (λ_GC_) were 1.10 (eGFRcr), 1.00 (eGFRcys), and 1.03 (sRAGE, Supplementary Table 2), which are acceptable for GWAS and indicate an absence of genomic inflation, systematic technical bias, or population stratification. The Manhattan plots (Supplementary Figure 2) represent the GWAS −log_10_ (*p*-value) on genomic scales for eGFRcr, eGFRcys, and sRAGE. We identified ten novel loci at GWAS *p*<5×10^-8^ (Table 1), including five loci for eGFRcr (1p32.3, *CDKN2C/MIR4421*; 4q28.3,

**Table 1.** Summary of novel locus variants identified for kidney function from correlated meta-analysis.

| Locus | Traits | SNP | Chr | Locus | eGFRcr<br>*p*-value | eGFRcys<br>*p*-value | sRAGE<br>*p*-value | CMA<br>*p*-value |
| --- | --- | --- | --- | --- | --- | --- | --- | --- |
| 1 | eGFRcr_eGFRcys | rs547464256 | 1p34.3 | *LINC01343/LINC01685* | 3.02x10<sup>-6</sup> | 1.68x10<sup>-6</sup> |  | 1.49x10<sup>-8</sup> |
| 2 | eGFRcr_eGFRcys_sRAGE | rs533133043 | 1p32.3 | *CDKN2C/MIR4421* | 5.02x10<sup>-8</sup> | 4.28x10<sup>-7</sup> | 4.08x10<sup>-4</sup> | 3.88x10<sup>-12</sup> |
| 3 | eGFRcr_sRAGE | rs6656882 | 1p22.3 | *SH3GLB1/SELENOF* | 2.34x10<sup>-4</sup> |  | 2.34x10<sup>-10</sup> | 1.02x10<sup>-11</sup> |
| 4 | eGFRcr_eGFRcys | rs74100345 | 1p22.2 | *ZNF326/SNORD3G* | 2.98x10<sup>-6</sup> | 4.74x10<sup>-6</sup> |  | 3.12x10<sup>-8</sup> |
| 5 | eGFRcr_eGFRcys_sRAGE | rs76181979 | 1p21.2 | *LINC01307* | 4.85x10<sup>-6</sup> | 6.28x10<sup>-5</sup> | 9.54x10<sup>-3</sup> | 4.74x10<sup>-8</sup> |
| 6 | eGFRcr_eGFRcys | rs115559549 | 1q24.1 | *LOC440700/TMCO1* | 2.20x10<sup>-6</sup> | 2.61x10<sup>-7</sup> |  | 3.19x10<sup>-9</sup> |
| 7 | eGFRcr_eGFRcys | rs139831128 | 2q37.2 | *ASB18/IQCA1* | 1.36x10<sup>-5</sup> | 9.67x10<sup>-7</sup> |  | 3.12x10<sup>-8</sup> |
| 8 | eGFRcr_eGFRcys | rs753994697 | 3p24.3 | *KCNH8* | 2.97x10<sup>-7</sup> | 6.04x10<sup>-5</sup> |  | 4.65x10<sup>-8</sup> |
| 9 | eGFRcr_eGFRcys | rs562793857 | 3p12.1 | *LINC02070/VGLL3* | 1.88x10<sup>-11</sup> | 3.76x10<sup>-9</sup> |  | 3.51x10<sup>-14</sup> |
| 10 | eGFRcr_eGFRcys | rs16850353 | 3q23 | *CLSTN2* | 1.62x10<sup>-7</sup> | 2.60x10<sup>-5</sup> |  | 1.58x10<sup>-8</sup> |
| 11 | eGFRcr_eGFRcys_sRAGE | rs151274284 | 4p16.2 | *STK32B* | 1.60x10<sup>-5</sup> | 1.93x10<sup>-7</sup> | 1.36x10<sup>-3</sup> | 3.71x10<sup>-10</sup> |
| 12 | eGFRcr_eGFRcys_sRAGE | rs749822346 | 4p15.31 | *LCORL/SLIT2* | 8.97x10<sup>-7</sup> | 1.52x10<sup>-4</sup> | 6.19x10<sup>-3</sup> | 2.13x10<sup>-8</sup> |
| 13 | eGFRcr_eGFRcys | rs554254460 | 4q28.3 | *PCDH10/PABPC4L* | 1.63x10<sup>-9</sup> | 5.58x10<sup>-6</sup> |  | 2.32x10<sup>-10</sup> |
| 14 | eGFRcr_eGFRcys | rs72715959 | 4q34.3 | *LINC01098/LINC00290* | 5.13x10<sup>-6</sup> | 1.62x10<sup>-8</sup> |  | 9.53x10<sup>-10</sup> |
| 15 | eGFRcr_eGFRcys | rs28003 | 5p15.2 | *CTNND2* | 4.69x10<sup>-6</sup> | 1.89x10<sup>-6</sup> |  | 2.24x10<sup>-8</sup> |
| 16 | eGFRcr_eGFRcys_sRAGE | rs562304789 | 5q14.1 | *TENT2* | 5.72x10<sup>-4</sup> | 7.93x10<sup>-6</sup> | 6.78x10<sup>-4</sup> | 2.94x10<sup>-8</sup> |
| 17 | eGFRcr_eGFRcys_sRAGE | rs185704381 | 5q22.3 | *YTHDC2/KCNN2* | 1.65x10<sup>-5</sup> | 1.28x10<sup>-4</sup> | 2.17x10<sup>-3</sup> | 4.03x10<sup>-8</sup> |
| 18 | eGFRcr_eGFRcys_sRAGE | rs189218418 | 7p15.1-p14.3 | *CREB5* | 9.66x10<sup>-5</sup> | 2.04x10<sup>-5</sup> | 1.45x10<sup>-3</sup> | 2.67x10<sup>-8</sup> |
| 19 | eGFRcys_sRAGE | rs149174297 | 7q35-q36.1 | *CNTNAP2* |  | 1.02x10<sup>-5</sup> | 3.46x10<sup>-5</sup> | 6.58x10<sup>-9</sup> |
| 20 | eGFRcr_eGFRcys_sRAGE | rs230222 | 9q22.31 | *CENPP/ECM2* | 2.75x10<sup>-5</sup> | 1.09x10<sup>-4</sup> | 2.50x10<sup>-4</sup> | 8.37x10<sup>-9</sup> |
| 21 | eGFRcr_eGFRcys | rs7038363 | 9q22.33 | *PRXL2C/LOC441455* | 1.30x10<sup>-7</sup> | 8.33x10<sup>-5</sup> |  | 3.59x10<sup>-8</sup> |
| 22 | eGFRcr_eGFRcys_sRAGE | rs974573268 | 10p14 | *LINC00707* | 1.11x10<sup>-5</sup> | 1.26x10<sup>-4</sup> | 2.02x10<sup>-3</sup> | 2.90x10<sup>-8</sup> |
| 23 | eGFRcys_sRAGE | rs10905638 | 10p14 | *LOC101928272/LINC02670* |  | 1.91x10<sup>-6</sup> | 4.11x10<sup>-4</sup> | 1.95x10<sup>-8</sup> |
| 24 | eGFRcr_eGFRcys | rs184854082 | 11p14.3-p14.2 | *LUZP2/ANO3* | 3.41x10<sup>-8</sup> | 1.53x10<sup>-5</sup> |  | 3.74x10<sup>-9</sup> |
| 25 | eGFRcr_eGFRcys | rs116560702 | 11q12.2 | *MS4A6E/MS4A7* | 2.40x10<sup>-9</sup> | 1.25x10<sup>-8</sup> |  | 2.51x10<sup>-12</sup> |
| 26 | eGFRcr_eGFRcys_sRAGE | rs140810086 | 11q14.1 | *DLG2* | 6.16x10<sup>-6</sup> | 8.00x10<sup>-7</sup> | 1.63x10<sup>-3</sup> | 5.41x10<sup>-10</sup> |
| 27 | eGFRcys_sRAGE | rs73535710 | 11q14.3 | *DISC1FP1* |  | 3.56x10<sup>-5</sup> | 7.25x10<sup>-5</sup> | 4.14x10<sup>-8</sup> |
| 28 | eGFRcr_eGFRcys_sRAGE | rs184549240 | 11q22.1 | *LINC02737/CNTN5* | 2.92x10<sup>-5</sup> | 6.64x10<sup>-7</sup> | 9.94x10<sup>-5</sup> | 1.23x10<sup>-10</sup> |
| 29 | eGFRcr_eGFRcys_sRAGE | rs529684126 | 11q25 | *LOC283177/LINC02714* | 4.06x10<sup>-6</sup> | 6.29x10<sup>-5</sup> | 8.70x10<sup>-3</sup> | 3.87x10<sup>-8</sup> |
| 30 | eGFRcys_sRAGE | rs2895135 | 12q24.33 | *RIMBP2* |  | 6.73x10<sup>-4</sup> | 2.53x10<sup>-6</sup> | 4.19x10<sup>-8</sup> |
| 31 | eGFRcr_eGFRcys_sRAGE | rs112909432 | 14q12 | *LOC728755/FOXG1-AS1* | 1.17x10<sup>-6</sup> | 2.16x10<sup>-4</sup> | 8.47x10<sup>-3</sup> | 4.39x10<sup>-8</sup> |
| 32 | eGFRcr_eGFRcys_sRAGE | rs7141142 | 14q13.1 | *NPAS3* | 1.03x10<sup>-4</sup> | 3.88x10<sup>-6</sup> | 3.77x10<sup>-3</sup> | 2.33x10<sup>-8</sup> |
| 33 | eGFRcr_eGFRcys | rs186101700 | 14q24.3 | *GPATCH2/ESRRB* | 1.29x10<sup>-7</sup> | 3.22x10<sup>-5</sup> |  | 1.62x10<sup>-8</sup> |
| 34 | eGFRcr_eGFRcys_sRAGE | rs147775587 | 14q31.3 | *LINC02301/SNORD3P3* | 6.88x10<sup>-5</sup> | 3.48x10<sup>-5</sup> | 3.13x10<sup>-4</sup> | 8.38x10<sup>-9</sup> |
| 35 | eGFRcr_eGFRcys | rs543176745 | 15q25.3 | *AGBL1* | 8.14x10<sup>-9</sup> | 9.12x10<sup>-5</sup> |  | 6.99x10<sup>-9</sup> |
| 36 | eGFRcr_eGFRcys_sRAGE | rs528757227 | 17q24.3 | *CASC17—ROCR* | 7.52x10<sup>-4</sup> | 3.24x10<sup>-4</sup> | 1.37x10<sup>-5</sup> | 2.90x10<sup>-8</sup> |
| 37 | eGFRcr_eGFRcys | rs113025648 | 17q25.3 | *LINC01987/LINC01973* | 5.77x10<sup>-6</sup> | 3.50x10<sup>-6</sup> |  | 4.04x10<sup>-8</sup> |
| 38 | eGFRcr_eGFRcys | rs146289239 | 18p11.23 | *PTPRM* | 4.63x10<sup>-8</sup> | 3.07x10<sup>-7</sup> |  | 2.18x10<sup>-10</sup> |
| 39 | eGFRcr_eGFRcys | rs9963912 | 18q21.1-q21.2 | *SKA1/MAPK4* | 6.89x10<sup>-8</sup> | 7.46x10<sup>-5</sup> |  | 2.19x10<sup>-8</sup> |
| 40 | eGFRcr_sRAGE | rs73030728 | 19q12 | *LOC100420587/LINC00906* | 9.68x10<sup>-5</sup> |  | 1.64x10<sup>-5</sup> | 2.82x10<sup>-8</sup> |
| 41 | eGFRcr_eGFRcys_sRAGE | rs112684971 | 21q21.1 | *MIR99AHG/LINC01549* | 5.24x10<sup>-6</sup> | 3.03x10<sup>-5</sup> | 2.49x10<sup>-4</sup> | 1.05x10<sup>-9</sup> |
| 42 | eGFRcr_eGFRcys | rs144142100 | 21q22.3 | *PTTG1IP/ITGB2* | 1.31x10<sup>-4</sup> | 4.70x10<sup>-8</sup> |  | 2.80x10<sup>-8</sup> |

Notes: The most significantly associated SNPs per locus within a 1Mb region; Chr= cytological chromosome; eGFRcr, eGFRcys, and sRAGE *p*-values= GWAS *p* values for eGFRcr, eGFRcys and sRAGE, respectively; and CMA *p*-value = correlated meta-analysis *p*-value.

*PCDH10/PABPC4L*; 11p14.3-p14.2, *LUZP2/ANO3*; 15q25.3, *AGBL1*; and 18p11.23, *PTPRM*), two loci for eGFRcys (4q34.3, *LINC01098/LINC00290* and 21q22.3, *PTTG1IP/ITGB2*), two loci for eGFRcr and eGFRcys (3p12.1, *LINC02070/VGLL3* and 11q12.2, *MS4A6E/MS4A7*), and one locus for sRAGE (1p22.3, *SH3GLB1/SELENOF*). Noteworthy, the deleterious minor allele frequencies (MAF) of WGS variants for the eight novel loci associated with eGFR traits were lower than 0.5% in the LLFS, as well in the 1000 Genomes Project and the NCBI ALlele Frequency Aggregator (ALFA). A 0.5% or higher MAF has been generally used in the genotype imputation of the GWAS for kidney function variant discoveries, which could contribute to unidentified loci from large GWAS meta-analyses.^14, 15^ In addition, the imputation of variants with small MAF (<0.5%) cannot be reliable in imputed genotypes; thus, using WGS in LLFS allowed more accurate estimates of association and discovery of novel loci.

Further, to investigate whether pleiotropic genetic variants share effects on eGFR traits and sRAGE, we employed CMA analyses on GWAS *p*-values. The Manhattan plots (Supplementary Figure 3) display the CMA GWAS −log_10_ (*p*-value) on genomic scales. The estimated genomic inflation factors (λ_GC_<1.1, Supplementary Table 2) for CMA analyses were corrected for tetrachoric correlations for non-independence among GWAS (Supplementary Table 3). We identified 59 pleiotropic loci, of which 42 are novel discoveries for eGFR. As mentioned, ten of the 42 novel loci were detected in the GWAS of eGFR (N=9) and sRAGE (N=1). Still, their genomewide significance levels increased by employing the CMA approach. Among the 42 CMA loci, 17 were from combined GWAS *p*-values from eGFRcr, eGFRcys, with sRAGE, 19 from eGFRcr with eGFRcys, two from eGFRcr with sRAGE, and four from eGFRcys with sRAGE. Table 1 provides the GWAS and CMA *p*-values for the 42 lead novel variants, and the summary statistics results for all novel significant variants are in Supplementary Table 4. Figure 1 displays the locuszoom plots for some chromosome regions (1p22.3, 4q34.3, 9q22.31, 12q24.33, and 18q21.1-q21.2), and the 42 locuszoom plots are in Supplementary Figure 4.

**Figure 1.**
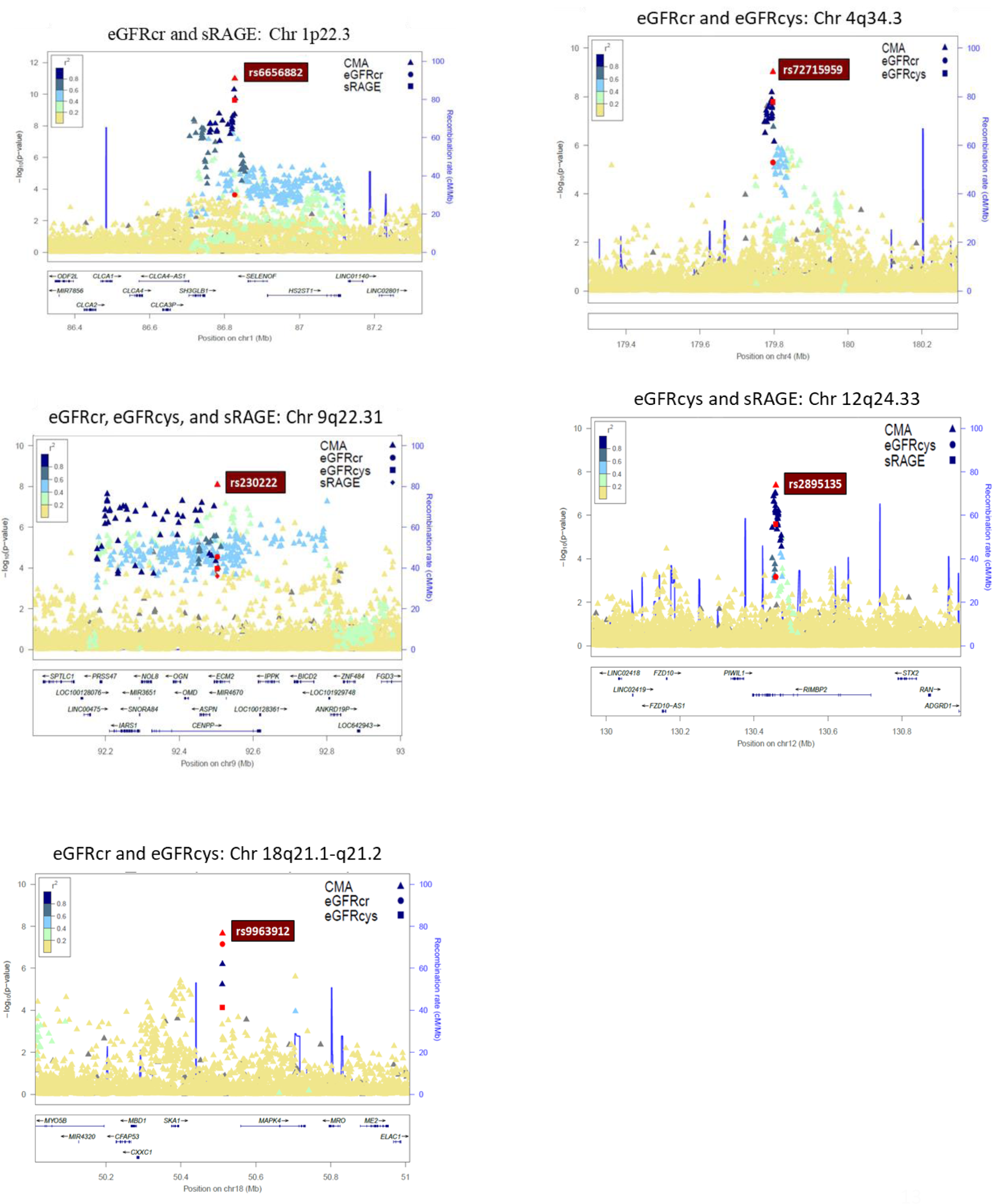
Locuszoom plots of correlated-meta analysis from GWAS for eGFRcr, eGFRcys, and sRAGE.

According to the GWAS catalog, 17 of 59 identified pleiotropic loci were reported for eGFR traits. Among the 17 loci, three loci for GWAS eGFRcr (*PROM1*, *AGPAT2*, and *KCQ2*) and one locus for GWAS eGFRcys (*USP32P1/KRT16P2*) reached genomewide association significance in the LLFS data (Supplementary Tables 5 and 6). For the other 13 loci that presented suggestive associations for kidney function in LLFS, the CMA approach enhanced the power of the joint association of eGFRcr, eGFRcys, and sRAGE, thereby replicating prior eGFR results from the literature.

### Functional annotations and regulatory elements from CMA GWAS

We searched the GWAS catalog to assess whether any of the 42 novel loci were identified in prior GWAS for kidney-related phenotypes and diseases but not with eGFRcr and eGFRcys in overall populations (Supplementary Table 4). Four out of 42 novel loci (1p32.3, *CDKN2C/MIR4421*; 1p22.2, *ZNF326/SNORD3G*; 4p16.2, *STK32B*; and 17q25.3, *LINC01987/LINC01973*) were reported at genomewide significance level for CKD, kidney cell carcinoma, and levels of blood urea nitrogen, serum uric acid, urate, serum creatinine, and creatine kinase. Four loci (4q34.3, *LINC01098/LINC00290*; 7q35-q36.1, *CNTNAP2*; 9q22.33, *PRXL2C/LOC441455*; and 18p11.23, *PTPRM*) showed suggestive associations (5×10^-8^<*p*<1×10^-5^) with CKD, eGFR in CKD, diabetic kidney disease, and uric acid. In addition, three loci (12q24.33, *RIMBP2*; 17q25.3, *LINC01987/LINC01973*; and 18q21.1-q21.2, *SKA1/MAPK4*) were associated with longevity. These findings show that some novel-identified loci for kidney function, previously described for kidney-related diseases from the literature, can be involved in biological pathways for developing kidney diseases. In contrast, other novel loci can predispose individuals to longevity.

We accessed the NCBI-dbSNP, HaploReg, GTEx-v8, and Human Kidney eQTL Atlas to annotate CMA GWAS variants (*p*<5×10^-8^) concerning their functional consequence and regulatory potential and TCGA database to verify whether the locus genes (harboring SNPs at *p*<5×10^-8^) predicted kidney carcinoma. The HaploReg tool indicated a total of 12 SNPs (5 loci) mapped in conserved syntenic regions by GERP or SiPhy, 26 SNPs (13 loci) associated with promoter histone marks, 84 SNPs (27 loci) with enhancer histone marks, 48 SNPs (21 loci) located at DNAse hypersensitive sites, 22 SNPs (16 loci) at protein regulatory binding sites, and 169 SNPs (41 loci) in transcription factor (TF) binding motifs (Supplementary Table 7). The GTEx-v8 project contained cis-eQTLs for 36 variants (three loci). Thirty-two variants on the *SH3GLB1/SELENOF* locus (1p22.3, Figure 1) were cis-eQTLs for *SH3GLB1* in skeletal muscle, and 19 variants were cis-eQTLs for *SELENOF* in ganglia brain, cortex brain, and thyroid (Supplementary Table 8). Three variants of *CENPP/ECM2* (9q22.31, Figure 1) were eQTLs for *OGN*, *CENPP*, lncRNA *RP11-526D8.11*, *NOL8*, ECM2, and *ANKRD19P* in several tissues, including whole blood, skeletal muscle, tibial nerve, brain, heart, artery, and adipose. One variant of *PRXL2C/LOC441455* (9q22.33) was an eQTL for *MFSD14C* in whole blood, skeletal muscle, artery, and adipose. According to the Human Kidney eQTL Atlas, 37 variants (two loci) were cis-eQTLs for target genes in the kidney.

Thirty-four variants in *SH3GLB1/SELENOF* were eQTLs for *SELENOF* (*SEP15*) and *HS2ST1* in the kidney glomeruli, and 15 were eQTLs in kidney tubules. Three variants in the *CENPP/ECM2* locus were eQTLs for *NOL8* and *CENPP* in kidney glomeruli and tubules (Supplementary Table 8). In addition, kidney eQTL meta-analysis from the Human Kidney eQTL Atlas reported 23 *SH3GLB1/SELENOF* variants as kidney eQTLs for *SELENOF* and three *CENPP/ECM2* variants as kidney eQTLs for *NOL8* and *CENPP*. Hence, these benchmark results suggest that several genetic variants in novel loci have functional regulatory effects, including glomerular filtration and tubular reabsorption in the kidney. In addition, *SH3GLB1*, *SELENOF,* and *ECM2*, among the other 25 locus genes, predicted kidney carcinoma in the TCGA database. The Venn diagram in Figure 2 represents the 42 novel CMA GWAS loci and the overlapped loci with HaploReg, GTEx-v8, Human Kidney eQTL Atlas, and TCGA databases.

**Figure 2.**
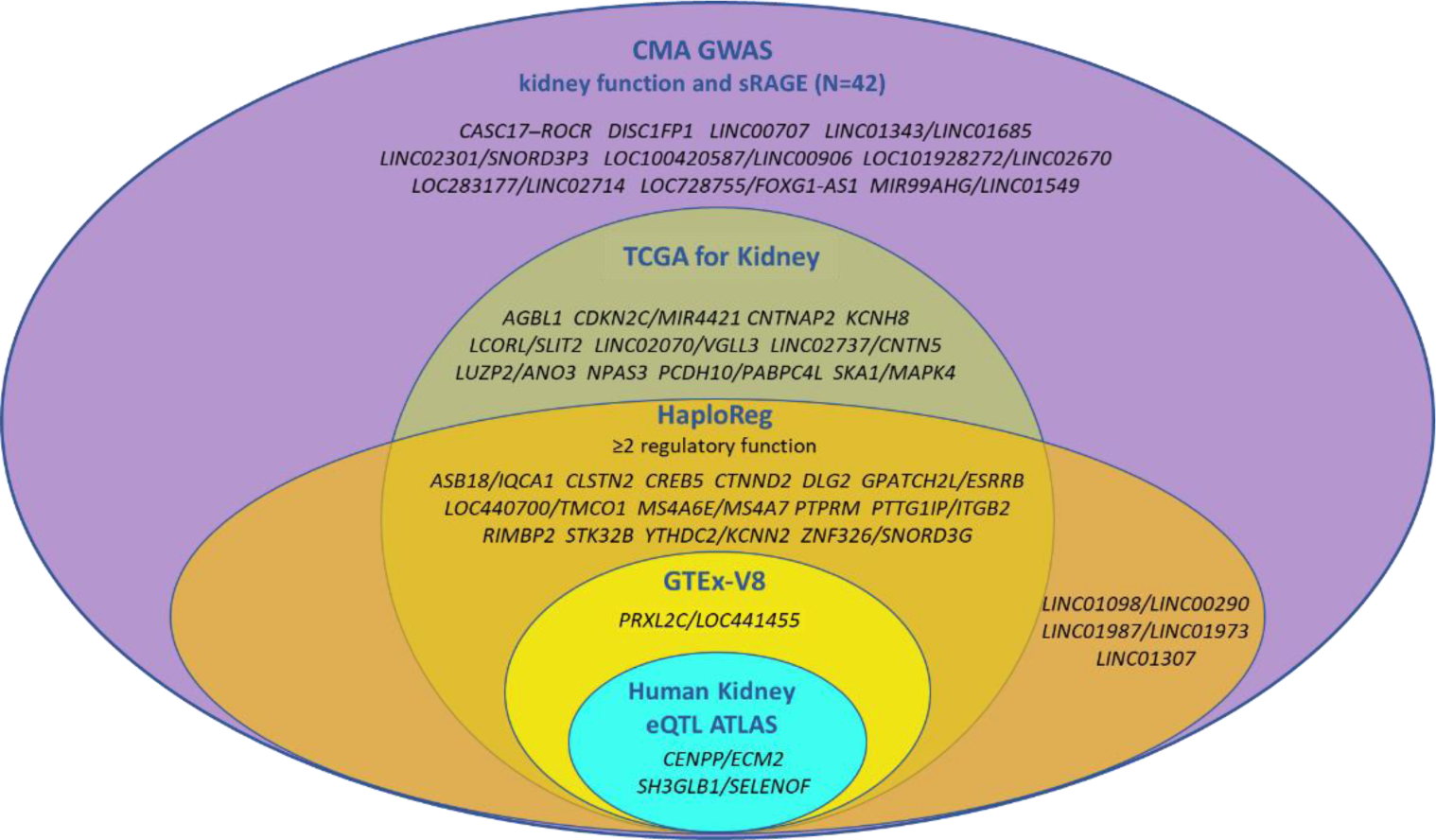
Venn diagrams for CMA GWAS loci overlapped with Human Kidney eQTL Atlas, GTEx, HaploReg, and TCGA.

### Transcriptome-wide association study and correlated meta-analysis

We performed TWAS analysis for eGFRcr, eGFRcys, and sRAGE on blood RNA-seq data to identify gene expression levels of protein-coding genes and lncRNAs. After applying the Bioconductor Bacon package to correct bias and inflation in TWAS, the genomic inflation factors were 1.03 (eGFRcr), 1.15 (eGFRcys), and 1.02 (sRAGE). Supplementary Figure 5 shows the observed versus expected TWAS −log_10_ (*p*-value) distributions for eGFRcr, eGFRcys, and sRAGE.

We found four genes with significant expression levels (*p*< 2.73×10^-6^) for kidney function from TWAS (Table 2), including *ODC1* for eGFRcr, *ERGIC1* and *LSP1* for eGFRcys, and *C16orf54* for eGFRcr and eGFRcys. In addition, CMA TWAS identified 14 gene expressions for kidney function, in which TWAS sRAGE contributed to discovering 9 out of 17 pleiotropic genes.

**Table 2.**
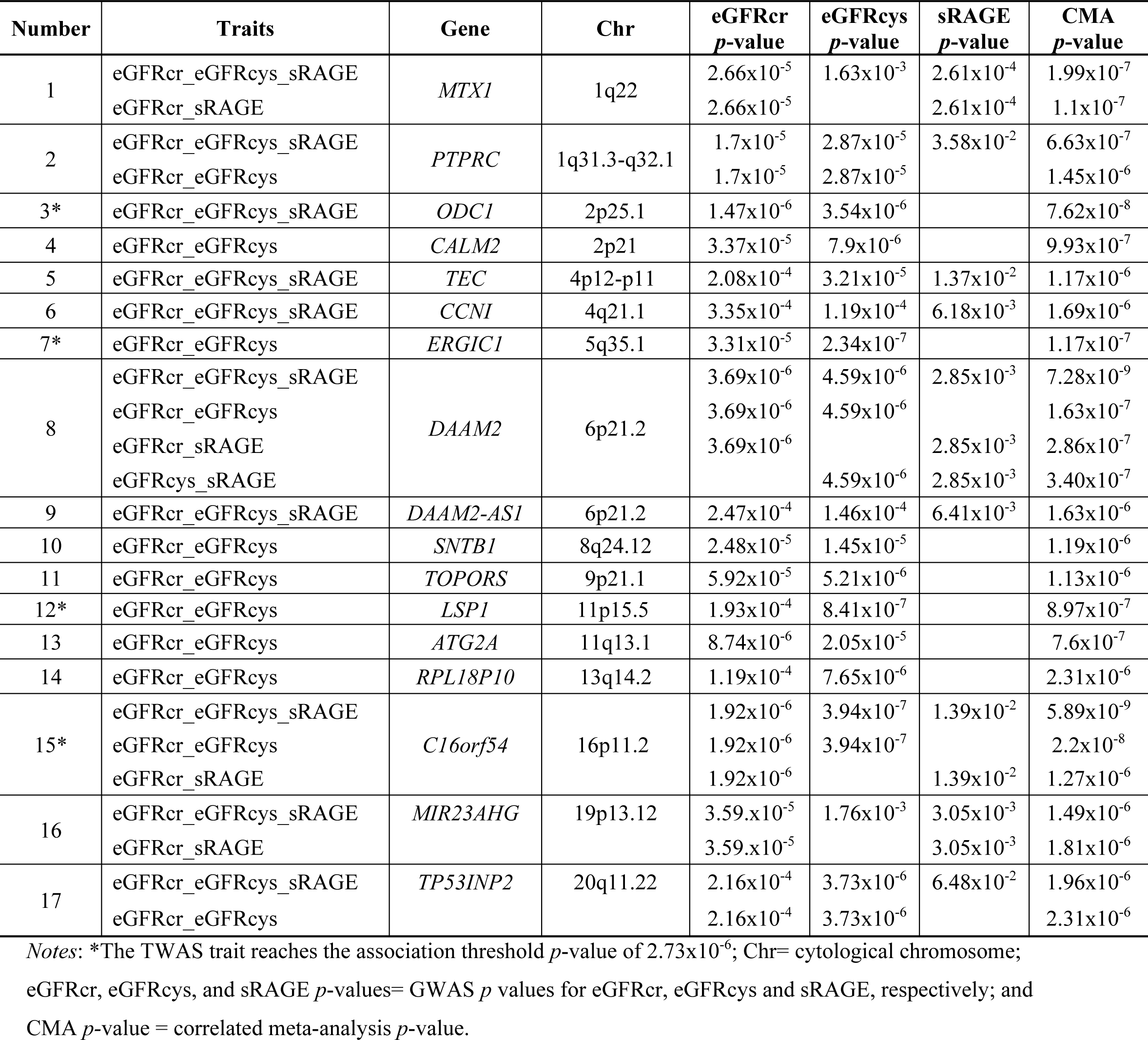
Summary of genes identified for kidney function from CMA TWAS.

### Regulatory elements from CMA TWAS

To explore our findings on the regulatory function from published eQTL, kidney carcinoma, and other cancer predictions, and enhancer-promoter informatively, we accessed the Human Kidney eQTL Atlas, GeneHancer database genomewide integration from the framework of GeneCards, TCGA, and EPRI maps. We found that 9 out of 17 CMA TWAS (*MTX1*, *PTPRC*, *CALM2*, *TEC*, *CCNI*, *DAAM2*, *TOPORS*, *LSP1*, and *ATG2A*) predicted kidney carcinoma in the TCGA database (Supplementary Table 9). The Human Kidney eQTL Atlas showed evidence that six genes harbored eQTLs expressed in kidney tissues. The eQTLs for *TEC*, *CCN1*, *ERGIC1*, *SNTB1*, and *LSP1* affected expression levels in kidney glomeruli, and *LSP1* and TOPORS had eQTLs for expression levels in the kidney tubules. In addition, eQTLs for *DAAM2-AS1*, *TOPORS,* and *LSP* were reported in the Human Kidney eQTL Atlas meta-analysis (Supplementary Table 9). The GeneHancer identifiers connected seven enhancer-promoter gene targets (*MTX1*, *CALM2*, *SNTB1*, *TOPORS*, *LSP1*, *ATG2A*, and *TP53INP2*) which were associated with kidney function-related phenotypes at genomewide significance in the GWAS catalog (Supplementary Table 10). The EPRI maps of cancer-associated variants in enhancer and promoter regions from the ICGC database (EPRI-ICGC) were associated with expressions of 12 genes (*MTX1*, *PTPRC*, *ODC1*, *CALM2*, *TEC*, *CCNI*, *ERGIC1*, *DAAM2*, *TOPORS*, *LSP1*, *ATG2A*, and *TP53INP2*, Supplement Table 11). A brief description of 17 genes identified by CMA TWAS is in Supplement Table 12, which shows that ten genes may play some role in the kidney. The Venn diagram in Figure 3 represents the genes identified by CMA TWAS and the overlap genes with CMA GWAS, kidney carcinoma from the TCGA database, and genes with regulatory elements affecting the kidney function from the Human Kidney eQTL Atlas, GeneHancer-GWAS catalog, and cancer-associated enhancer-promoter variants from the EPRI-ICGC.

**Figure 3.**
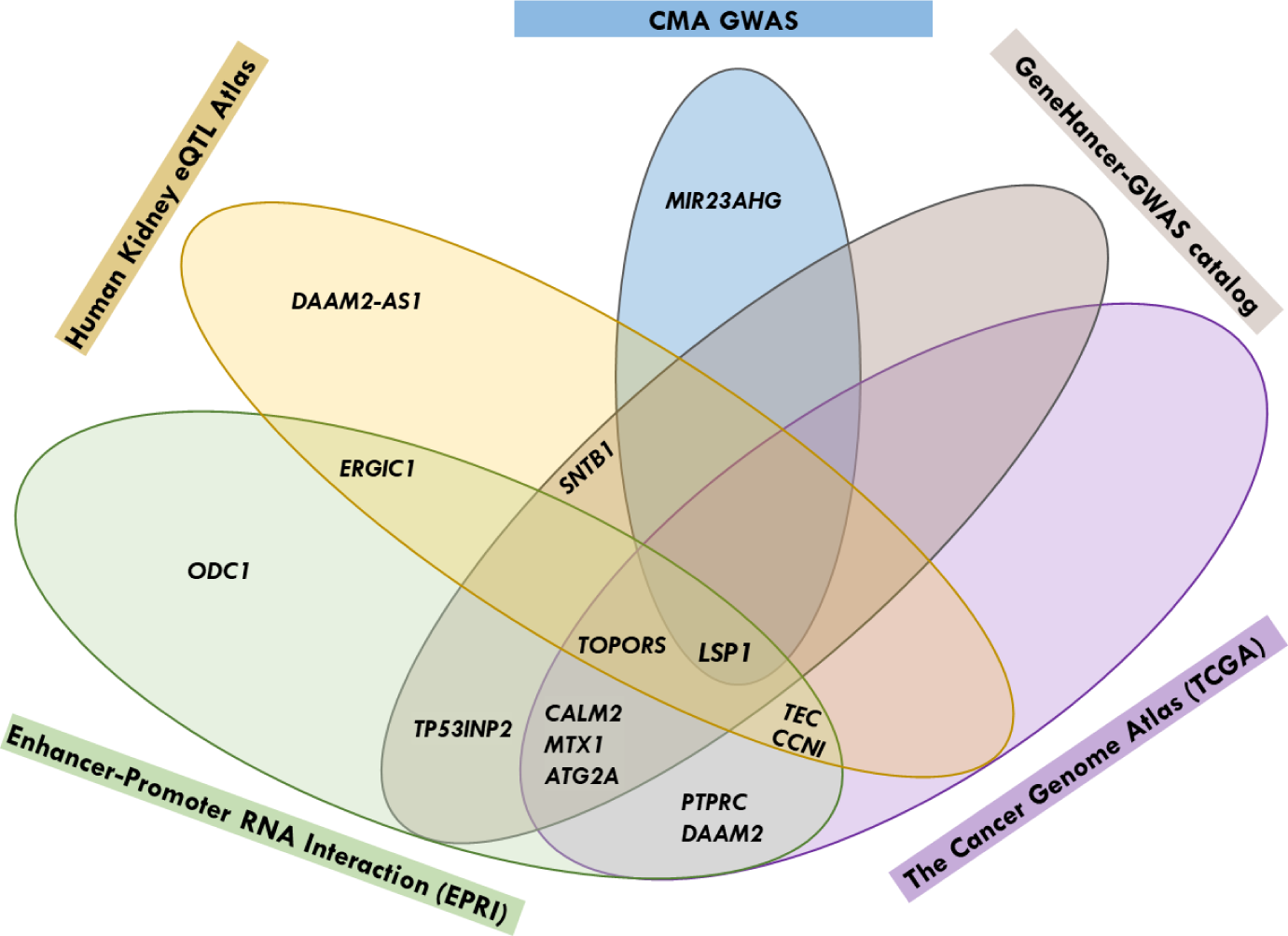
Venn diagrams for CMA TWAS genes overlapped with CMA GWAS, Human Kidney eQTL Atlas, TCGA, EPRI, and GeneHancer-GWAS Catalog.

Of note, three EPRI-ICGC variants in the enhancer region predicting *LSP1* expressions are located at ∼380 Kb upstream of GWAS *DLG2*-intronic rs140810086 (11q14.1, Supplementary Table 4), and the *LSP1* gene is at ∼230 Kb downstream of GWAS lncRNA-*KCNQ1OT1* exonic-rs2157899 (11p15.5, Supplementary Table 6). Figure 4 represents the long-range enhancer-promoter looping between EPRI-ICGC enhancer variants with *LSP1* expression. Four EPRI-ICGC variants in enhancer regions predicting *MTX1* and *MTX1LP* gene expressions were also associated with eGFR, serum creatinine levels, blood urea nitrogen levels, urea levels, urate levels, serum uric acid levels, and gout, as reported in the GWAS catalog (Supplement Table 11). In addition, the *MIR23AHG* gene identified by CMA TWAS (Table 2) at 19p13.12 is ∼100 Kb from the GWAS *CACNA1A/ CCDC130* intergenic-rs563793231, which was previously associated with kidney function in the GWAS catalog.

**Figure 4.**
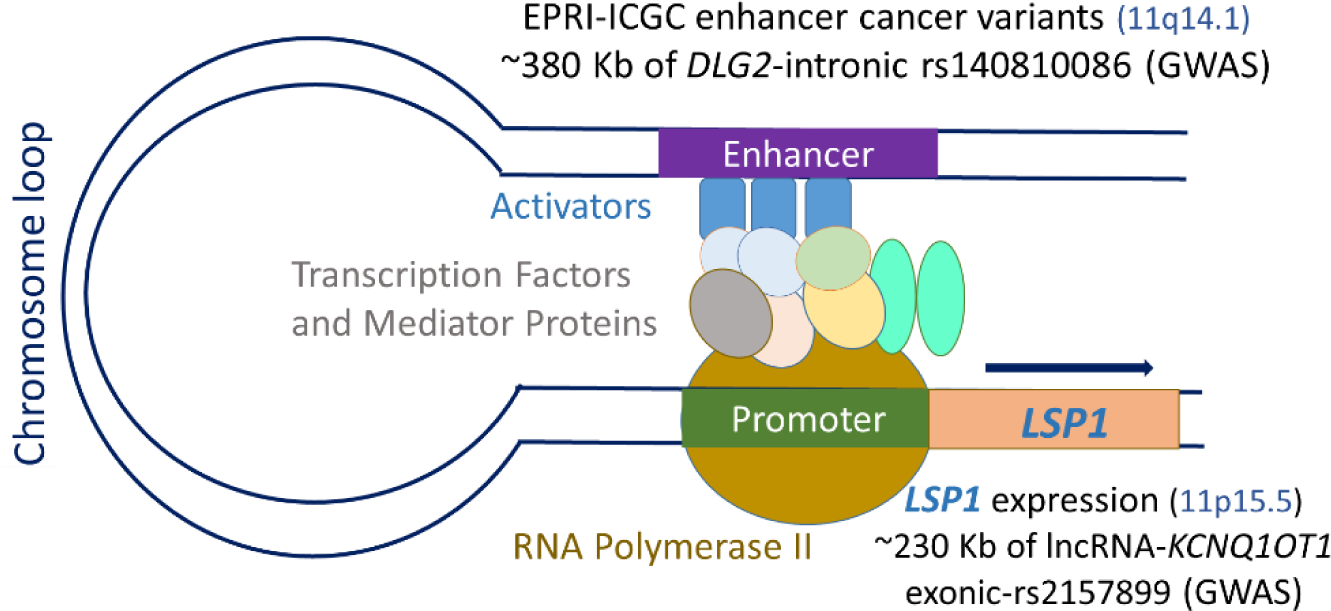
Diagram of EPRI maps of cancer-associated enhancer-variants from the ICGC database (EPRI-ICGC) predicting *LSP1* expression.

## Discussion

The prevalence of CKD (28.3%) in healthy-aging LLFS, at the average age of 70.4 years, is lower than the overall US population aged 65 years or older (34% measured by eGFRcr).^26^ LLFS individuals presented better kidney function measured by lower creatinine levels when compared to similarly aged individuals in CHS (Cardiovascular Health Study) and FHS (Framingham Heart Study).^20^ Low creatinine levels were also associated with becoming a centenarian in exceptionally long-lived individuals in the Swedish AMORIS (Apolipoprotein-related MOrtality RISk) cohort.^27^

The mean eGFR measured by cystatin C was higher than that measured by creatinine in LLFS (eGFRdiffcys-cr= eGFRcys – eGFRcr*=*5.4±17.5 ml/min/1.73 m^2^), which is in agreement with other studies in healthy older people. Positive differences of eGFRdiffcys-cr were associated with lower comorbidities and better functional status in HABC (Health, Aging, and Body Composition Study),^7^ lower risks for adverse longitudinal outcomes and mortality in SPRINT (Systolic Blood Pressure Intervention Trial),^8^ and lower risk for incident frailty and mortality in CHS.^9^ Serum cystatin C level is a biomarker of kidney aging,^28^ and their higher levels have been associated with poor physical function and cognition^7^. In contrast, decreased muscle mass in the aged may mask low creatinine levels. Although estimates of GFR by creatinine and cystatin C levels may lack precision and accuracy in clinical practice, early kidney disease diagnosis can be improved using both eGFRcr and eGFRcys, as recommended by the KDIGO.^12^

### Biological Insights from CMA GWAS

eGFRcr, GFRcys, and sRAGE are correlated, indicating shared genetic effects and biological pathways. The power gained by accounting for the GWAS correlations between eGFRcr with eGFRcys and with sRAGE via the CMA approach enabled the discovery of 59 loci for eGFR traits, including 42 novel and 17 previously reported in the GWAS catalog for eGFR. Eleven of the 42 novel loci, which were previously associated with kidney-related traits (1p32.3, 1p22.2, 4p16.2, 4q34.3, 7q35-q36.1, 9q22.33, 17q25.3, and 18p11.23, Supplementary Table 4) or longevity (12q24.33, 17q25.3, and 18q21.1-q21.2) in the GWAS catalog but not with eGFRcr and eGFRcys, predict kidney diseases and kidney anomalies. The findings suggest that kidney-related loci may be involved in reduced GFR and progressive glomerular, tubular, and interstitial damage. Moreover, several loci harbor genes that participate in apoptosis, inflammation nuclear factor (NF)-κB, Wnt/β-catenin, transforming growth factor (TGF)-β signaling pathways, mineralocorticoid receptor-mediated transactivation, and ubiquitin-dependent processes (as summarized in the Appendix, Supplementary Materials). On the other hand, longevity-related loci are related to human synaptic development, function, plasticity, and kidney-related diseases.

Noteworthy, variants from four pleiotropic loci (17q24.3, 13q12.2, 9q22.31, and 1p22.3) had protective effects for eGFR traits in LLFS. At 17q24.3 locus, the *CASC17/ROCR* rs528757227-A frequency in LLFS (0.0131, Supplementary Table 4) was ∼5.34 and ∼2.34 times higher than in ALFA-Europeans (0.0025) and TOPMed (0.0056), respectively. The intergenic-rs528757227 variant is located within two lncRNA genes, *CASC17* (cancer susceptibility 17) and *ROCR* (regulator of chondrogenesis RNA), which have unknown roles in the kidney.

At 13q12.2 locus, the *POLR1D/GSX1* rs56058022-C allele frequency was ∼1.51 and ∼1.87 times higher in LLFS (0.0257, Supplementary Table 5) than in ALFA-Europeans (0.0170) and TOPMed (0.0138), respectively. The rs56058022 is at 30 Kb 5’ of *GSX1* (GS Homeobox 1), which acts upstream of or within positive regulation of transcription by RNA polymerase II. Kidney function association at *POLR1D/GSX1* locus was previously described between *FLT3* (∼310-Kb downstream of rs56058022, Supplementary Table 6) with eGFRcr.

At 9q22.31 locus (Figure 1), *CENPP/ECM2* rs230222-G had a slightly higher allele frequency in LLFS (0.4887) than in ALFA-Europeans (0.4330) and TOPMed (0.3982). The rs230222 was a QTL for *CENPP* in the kidney tubules (Supplementary Table 8). *CENPP* (centromere protein P) assembles kinetochore proteins, mitotic progression, and chromosome segregation. *CENPP* was previously associated with urate levels and CKD. In a mouse model, a transcriptomics analysis demonstrated that the *cenpp* belonged to a small population (<1%) in proliferation-related genes with higher expression of cell-cycle machinery with unique attributes of distal convoluted tubule cells.^29^ *CENPP* rs230222 was also a QTL for *NOL8* in the kidney glomeruli and tubules. *NOL8* predicted predicted kidney carcinoma in the TCGA database. Although the genes *CENPP* and *NOL8* were connected with kidney regulation, their precise role in kidney physiology still needs to be understood. These relatively higher allele frequencies of protective variants in *CASC17/ROCR*, *POLR1D/GSX1*, and *CENPP/ECM2* reflect better kidney function in the health-aging LLFS members than in general populations.

The variants of *SH3GLB1/SELENOF* (1p22.3, Figure 1) had protective effects for the kidney, but their common allele frequencies were lower in LLFS (e.g., rs6656882-C =0.2052) than in the ALFA-Europeans (0.2764) and TOPMed (0.3461). The Human Kidney eQTL Atlas reported that the *SH3GLB1/SELENOF* intergenic rs6656882 was a cis-eQTL for *HS2ST1* in the kidney glomeruli and an eQTL for *SELENOF* in the kidney glomeruli and tubules (Supplementary Table 8). *SELENOF* encodes a protein of the SEP15/selenoprotein M family that is the primary mediator of selenium effects in human health. *HS2ST1* encodes the heparan sulfate 2-O-sulfotransferase 1, which belongs to the group of glycosaminoglycans involved in multiple signaling pathways. Kidney agenesis was described in the *Hs2st1*−/− mutant embryonic mouse^30^ and in the human *HS2ST1* bi-allelic pathogenic variants,^31^ which suggests that the absence of heparan sulfate interferes with the signaling required for kidney formation. TCGA database also shows that *SH3GLB1*, *SELENOF,* and *HS2ST1* predicted kidney carcinoma; however, their roles in kidney dysfunction remain unclear.

### Biological Insights from CMA TWAS

Among 17 genes identified by CMA TWAS, 15 gene expressions were: (i) within CMA GWAS loci (2 genes), (ii) predictive of kidney carcinoma (TCGA, 9 genes), (iii) harboring eQTLs in the kidney glomeruli and tubules (Human Kidney eQTL Atlas, 9 genes), (iv) associated with kidney function-related phenotypes at genomewide significance using enhancer–targeted gene data (GeneHancer-GWAS catalog, 7 genes), or (v) cancer-associated variants in enhancer-promoter regions predicting gene expressions (EPRI-ICGC, 12 genes). *LSP1* was present in the five data sources, highlighting it as a prominent gene for further investigation into kidney function-related phenotypes (Figure 3). *LSP1* regulates the actin cytoskeleton’s structural organization and was reported to be a WT1 target gene expressed during kidney development.^32^ It is worth mentioning that *DLG2* and lncRNA-*KCNQ1OT1* GWAS loci may participate in *LSP1* expression. Three cancer-associated variants in the enhancer region from the EPRI-ICGC database (11q14.1) predicting *LSP1* (11p15.5) expressions are located at ∼380 Kb of *DLG2*-intronic rs140810086 (11q14.1), while the *LPS1* gene is located at ∼230 Kb of lncRNA-*KCNQ1OT1* exonic-rs2157899 (11p15.5). These findings suggest a long-range enhancer-promoter looping between EPRI-ICGC enhancer variants with *LSP1* expression (Figure 4). The *DLG2*-intronic rs140810086 may be part of the enhancer region in high LD with the EPRI-ICGC variants predicting *LSP1* expression. The lncRNA-*KCNQ1OT1* is an antisense of *KCNQ1*, which was previously associated with kidney function and related phenotypes in the GWAS catalog. *KCNQ1OT1* is implicated in epigenetic gene silencing in imprinting, affecting gene expressions and various cell functions, such as cell proliferation, migration, apoptosis, and inflammation.^33^ *KCNQ1OT1* interference reduced the expression of inflammatory factors in high glucose-induced HK-2 (derived Human Kidney) and decreased oxidative stress and pyroptosis of kidney tubular epithelial cells in diabetic nephropathy patients.^34^ In acute kidney injury (AKI) knockdown mouse, *Kcnq1ot1* promoted miR-204-5p expression, inhibited *NLRP3* inflammasome activation, reduced levels of serum creatinine, blood urea nitrogen, and kidney injury molecule-1, and thus alleviated AKI and reduced apoptosis.^35^ However, to confirm whether *LSP1* expression is regulated by GWAS locus variant at 11q14.1 (such as *DLG2*-intronic rs140810086) and epigenetically regulated by lncRNA-*KCNQ1OT1* at 11p15.5, further investigations of molecular mechanisms are required.

*MIR23AHG*, *TOPORS*, and *TP53INP2* are prominent genes for exploration into kidney disease, among the other 15 gene expressions presented in two or more data sources. The lncRNA-*MIR23AHG* (mir-23a/27a/24-2 cluster) identified by CMA TWAS (Table 2, Figure 3) at 19p13.12 is ∼100 Kb from the *CACNA1A/CCDC130* intergenic-rs563793231 that showed association with CMA GWAS for eGFRcr, eGFRcys, and sRAGE. The miR-23a-3p regulated the inflammatory response and fibrosis, attenuating the development of diabetic kidney disease in mice through the *Egr1* gene, which has a crucial role in renal tubular injury.^36^ Another mouse study demonstrated that miR-23a-3p ameliorates sepsis-induced AKI by targeting *FKBP5* and inactivating the NF-κB signaling.^37^

*TOPORS* showed evidence of involvement in kidney function-related phenotypes in the four data sources: TCGA, Human kidney eQTL Atlas, GeneHancer-GWAS, and EPRI (Figure 3). *TOPORS* is a tumor suppressor involved in cell growth, cell proliferation, and apoptosis that regulates p53/TP53 stability through ubiquitin-dependent degradation.^38^ *TOPORS* mRNA expression was lower in stages III and IV than in the earlier stages of kidney carcinoma (KIRC). Patients in the low levels group presented shorter survival than those with high levels of *TOPORS* in KIRC.^39^

*TP53INP2* was identified for predicting kidney-related diseases in GeneHancer-GWAS and EPRI (Figure 3). *TP53INP2* (tumor protein p53-inducible nuclear protein 2) was significantly lower in renal cell carcinoma (RCC) than in normal kidney cells. Overexpressed *TP53INP2* suppressed the activity, migration, and invasion of RCC cells, inhibiting mouse tumor growth and promoting cell apoptosis. T*P53INP2* induced apoptosis in RCC cells through the caspase-8/TRAF6 pathway.^40^

### Strengths and limitations

Our study has important strengths and some limitations. Several GWAS variants/genes and TWAS genes have confirmed evidence of gene expressions in kidney glomerular, kidney tubular, kidney carcinoma, and biological pathways impacting kidney function-related phenotypes. However, statistically significant GWAS and TWAS results can suggest association but not causation, and further molecular studies are needed. The GWAS variants do not necessarily reside in the genes’ proximity. They can modulate transcriptional activities of target genes as far away as a cis-eQTL (∼1 Mb of a gene) or up to several megabases away as a trans-eQTL (>5 Mb away from a gene or on another chromosome).^41^ Additionally, TWAS is a gene-based association in which eQTLs jointly regulate the transcriptional activities of a gene, but eQTLs explain only a tiny fraction of the GWAS signals. GWAS and cis-eQTL hits are systematically different due to bias toward more constrained genes, transcription factors, and complex regulatory landscapes in GWAS, while eQTLs show a strong promoter bias.^42^ Other restraints that could limit the power of TWAS are the relatively small subset of LLFS individuals (N=1,209) available for the current study and the gene expression levels were measured on whole blood RNA-seq, not specifically in kidney tissue. In addition, the generalization of our findings to other populations and ancestry groups needs confirmation because the allele frequency of the associated variants and the LD in European ancestry can differ among populations.

## Conclusion

The current study reveals that assessing genome and transcriptome data from the healthy-aging LLFS population helped capture the complexity of biological regulatory mechanisms in kidney function and can prioritize chromosomal regions for further genetic investigations. Several identified genes and gene expressions have potentially functional genome elements that can be implicated in pathways for kidney and healthy-aging biological pathways. The pleiotropy of circulating sRAGE levels and kidney function may shed novel insight for future kidney disease research.

## Data Availability

All data produced are available online at dbGaP (study accession: phs000397.v1.p1).

## Additional Information

Supplementary Material is available online.

## Acknowledgments

This work was supported by the National Institute on Aging (UI9AG023122 and U19AG063893). We thank the Long Life Family Study participants and its members.

## Author contributions

Conceptualization: M.F.F., M.A.P.; correlated meta-analysis method: M.A.P.; investigation (acquisition of clinical data): B.T., K.C., J.M.Z.; data curation: M.F.F., S.J.L., M.K.W., and M.A.P; performed GWAS, TWAS, and the statistical and bioinformatic analyses: M.F.F., S.J.L., S.A., and M.R.B.; writing: M.F.F.; review, edit, and approval of the manuscript: all authors.

## Declaration of Interest

The authors do not have any conflicts of interest to declare.

## Notes

### Competing Interest Statement

The authors have declared no competing interest.

