## Supplementary Materials for "Discovery of genomic and transcriptomic pleiotropy between kidney function and soluble receptor for advanced glycation end-products using correlated meta-analyses: The Long Life Family Study (LLFS)"

Feitosa MF et al.

**SUPPLEMENTARY MATERIAL**

**Appendix.** Summary of novel loci associated with kidney-related traits or longevity (docx)

**Supplementary Tables**

**Supplementary Table 1.** Characteristics of participants in the analyses (docx)

**Supplementary Table 2.** Genomic control (λ) of GWAS and CMA for kidney function (eGFRcr and eGFRcys) and sRAGE (docx)

**Supplementary Table 3.** Tetrachoric correlations of CMA for kidney function traits and sRAGE (docx)

**Supplementary Table 4.** Novel loci from CMA GWAS for kidney function, GWAS catalog, TCGA (xlsx)

**Supplementary Table 5.** Reported loci for kidney function and replicated by CMA GWAS (xlsx)

**Supplementary Table 6.** GWAS catalog for kidney function reported loci and replicated by CMA GWAS (xlsx)

**Supplementary Table 7.** HaploReg regulatory features for locus variants identified by CMA GWAS (xlsx)

**Supplementary Table 8.** GTEx and Human Kidney eQTL Atlas (xlsx)

**Supplementary Table 9.** CMA TWAS results, Human Kidney eQTL Atlas, and TCGA (xlsx)

**Supplementary Table 10.** CMA TWAS, GeneHancer-GeneCards, and GWAS Catalog (xlsx)

**Supplementary Table 11.** EPRI-ICGC, GWAS catalog, and CMA GWAS (xlsx)

**Supplementary Table 12.** Summary of literature for genes identified by CMA TWAS (xlsx)

**Supplementary Figures**

**Supplementary Figure 1.** GWAS quantile-quantile plots of observed versus expected $-\log_{10}$ (*p*-value)

**Supplementary Figure 2.** Manhattan plots of univariate GWAS for kidney function traits and sRAGE

**Supplementary Figure 3.** Manhattan plots of CMA GWAS for kidney function traits and sRAGE

**Supplementary Figure 4.** Locuszoom plots of novel CMA GWAS

**Supplementary Figure 5.** TWAS quantile-quantile plots of observed versus expected $-\log_{10}$ (*p*-value)

**Appendix.** Summary of novel loci associated with kidney-related traits or longevity

Correlated meta-analyses (CMA) on genome-wide association study (GWAS) *p*-values enable the identification of 42 novel pleiotropic loci for kidney function and sRAGE. Eleven of the 42 novel loci, which were previously associated with kidney-related traits (1p32.3, 1p22.2, 4p16.2, 4q34.3, 7q35-q36.1, 9q22.33, 17q25.3, and 18p11.23, Supplementary Table 4) or longevity (12q24.33, 17q25.3, and 18q21.1-q21.2) in the GWAS catalog but not with eGFRcr and eGFRcys, were involved in kidney diseases, kidney anomalies, and aging- and kidney-related signaling pathways.

Associations on chromosome 1p32.3 were reported between *CDKN2C* and *FAF1* with renal cell carcinoma, blood urea nitrogen levels, and serum uric acid levels. *CDKN2C* and *FAF1* predicted the prognosis of KIRC and KIRP, according to the TCGA database. *FAF1* was also involved in mediating apoptosis, nuclear factor (NF)-κB, Wnt/β-catenin and transforming growth factor (TGF)-β signaling pathways, mineralocorticoid receptor-mediated transactivation, and ubiquitin-dependent processes [1].

At 1p22.2 locus, *LRRC8C* was associated with serum uric acid levels. The LRRC8C protein is a critical component of T cells' volume-regulated anion channel (VRAC). The VRAC/LRRC8C suppresses T cell function, controlling T cell-mediated immune response by regulating cyclic dinucleotide transport and STING (stimulator of IFN genes)–p53 signaling [2]. VRAC/LRRC8 channels are also crucial for the function and integrity of proximal tubules. In mouse models, *lrrc8d* was prominently expressed in renal vascular endothelial cells, and its protein was colocalized with the endothelial marker ICAM1 in the outer medulla and glomerulus [3]. *LRRC8D* also predicted KIRC and KIRP in the TCGA database.

At 17q25.3 locus, associations were previously identified between *SOCS3/LINC01993* variants with creatine kinase levels and *TMC8* with serum creatinine and uric acid levels. *TMC8* is downstream regulated by microRNA-144-5p/oncogenic syndecan-3 axes associated with a poor prognosis of renal clear cell carcinoma (RCC) [4]. The other two-locus genes at 17q25.3, *TK1* and *BIRC5*, are biomarkers for predicting RCC prognosis [5, 6]. *TMC8*, *TK1*, and *BIRC5* also predicted poor RCC prognosis according to the TCGA database.

At the 4q34.3 locus, an intergenic variant was associated with the uric acid elevation in response to the thiazide-like diuretic in hypertension. The CMA GWAS for eGFRcr and eGFRcys identified 16 SNPs within a gene desert region on chromosome 4q34.3 (Figure 1, Supplementary Table 4). Approximately 1,240 Kb downstream of the lead SNP rs72715959 (MAF=0.0729) resides the closest gene, the lncRNA-240 (*LINC00290*), and at ~596 Kb downstream of rs777345385 (MAF=0.0029) locates the lncRNA-1098 (*LINC01098*). The terminal deletion on the 4q chromosome leads to a recognizable syndrome, including 4q34.3 deletion, with evidence of kidney anomaly [7] and autoimmune nephropathy [8].

At 9q22.33, an intergenic variant showed a suggestive association with eGFR in CKD patients. *CDC14B* ( 9q22.33) was strongly expressed in the apical proximal tubules in the nonneoplastic tissues, but its expression was completely absent in RCC cases [9]. The protein encoded by *CDC14B* antagonizes *CDK1*-mediated activating mitotic phosphorylation of the deubiquitinase *USP9X*, which targeted the Wilms' tumor protein-1 (WT1) [10]. The mutation of transcription factor *WT1* was reported to contribute to ~15% of aggressive pediatric kidney cancer [11].

In addition, associations were described between *STK32B* (4p16.2) with urate levels, *CNTNAP2* (7q35-q36.1) with diabetic kidney disease, and *PTPRM* (18p11.23) with dialysis survival [2]. The *STK32B*, *CNTNAP2*, and *PTPRM* genes predicted KIRC, and *PTPRM* also predicted KIRP in the TCGA database; however, the mechanisms involving *STK32B*, *CNTNAP2*, and *PTPRM* with kidney disease are unknown.

Moreover, three loci (12q24.33, 17q25.3, and 18q21.1-q21.2) identified from CMA GWAS for kidney function were also associated with longevity in the GWAS catalog (Supplementary Table 4). At 12q24.33 (Figure 1), the *RIMBP2* gene encodes a presynaptic protein involved in synaptic transmitter release at central synapses and also predicted KIRP and KIRC in the TCGA database. Mutations in *TCF4* were linked to dysregulation of *RIMBP2,* provoking several neurodevelopmental diseases and disrupting synaptic function in patient-derived cortical neurons, such as Pitt-Hopkins syndrome [12]. *TCF4* is a vital transcriptional regulator of human synaptic development, function, and plasticity [12]. It can also increase the severity of renal injury and contribute to the apoptosis of NRK-52E renal proximal tubular epithelial cells [13]. However, whether *RIMBP2* participates with *TCF4* in kidney diseases is unknown.

*SKA1* (18q21.1-q21.2, Figure 1) belongs to a microtubule-binding subcomplex of the outer kinetochore, which is essential for proper chromosome segregation and is involved in the growth and proliferation of numerous cancer types, including the pathogenesis of renal cell carcinoma [14]. *SKA1* also predicted KIRC in the TCGA database. *LINC01987/LINC01973* (17q25.3) locus was also significantly associated with kidney-related traits and longevity. Some genes in 17q25.3, such as *BIRC5* and *TK1*, predicted kidney carcinoma in the TCGA database, but their roles remain unclear.

**Supplementary Table 1.** Characteristics of participants in the analyses

| Variables | Mean (SD), or median (Q1, Q3) * or percentage |
|---|---|
| Number | 4182 |
| Age (years), [range] | 70.4 (15.7), [24 - 110] |
| Sex (male) | 45% |
| Hypertension | 52% |
| T2D | 6% |
| CHD | 9% |
| Serum sRAGE (pg/mL) | 631.21 ± 506.77<br>517.00 (371.00, 728.00) * |
| Serum Creatinine (mg/dL) | 1.05 ± 0.33<br>1.00 (0.87, 1.17) * |
| eGFRcr (ml/min/1.73 $m^2$) | 69.78 ± 18.42<br>70.65 (58.03,82.53) * |
| Serum Cystatin C (mg/dL) | 1.08 ± 0.43<br>0.95 (0.81, 1.20) |
| eGFRcys (ml/min/1.73 $m^2$) | 75.15 ± 27.15<br>78.54 (53.61, 98.40) |
| CKDcr cases (N, %) | 1182 (28.26%) |
| CKDcys cases (N, %) | 1286 (30.75%) |

*Notes*: mean (SD) = mean levels (standard deviation); median (first (Q1) and third (Q3) quartiles); hypertension = blood pressure (BP) above 140/90 mm Hg and/or taking anti-hypertensive medications; T2D = Type 2 diabetes; CHD = coronary heart disease; sRAGE = serum levels of soluble receptor for advanced glycation end products; eGFR = estimated glomerular filtration rate (eGFR), eGFRcr = eGFR from serum creatinine, eGFRcys = eGFR from serum cystatin C; CKDcr = chronic kidney disease (CKD) defined as an eGFRcr below 60 ml/min/1.73 $m^2$; CKDcys = CKD defined as an eGFRcys below 60 ml/mi/1.73 $m^2$. The mean ± SD of the difference of eGFRcys – eGFRcr: 5.37 ± 17.46 ml/min/1.73 $m^2$.

**Supplementary Table 2.** Genomic control (λ) of GWAS and CMA for kidney function (eGFRcr and eGFRcys) and sRAGE

| | eGFRcr | eGFRcys | sRAGE |
|---|---|---|---|
| GWAS / CMA | 1.106 | 0.996 | 1.030 |
| eGFRcr_eGFRcys_sRAGE | | | 1.094 |
| eGFRcr_eGFRcys | | 1.095 | |
| eGFRcr_sRAGE | | | 1.094 |
| eGFRcys_sRAGE | | | 1.094 |

*Notes*: eGFR = estimated glomerular filtration rate (eGFR), eGFRcr = eGFR from serum creatinine, eGFRcys = eGFR from serum cystatin C, sRAGE = serum levels of soluble receptor for advanced glycation end products.

**Supplementary Table 3.** Tetrachoric correlations of CMA for kidney function traits and sRAGE

| Traits | eGFRcr | eGFRcys | eGFRcr, eGFRcys |
|---|---|---|---|
| eGFRcys | 0.012 | | |
| sRAGE | 0.012 | 0.016 | 0.016 |

*Notes*: eGFR = estimated glomerular filtration rate (eGFR), eGFRcr = eGFR from serum creatinine, eGFRcys = eGFR from serum cystatin C, sRAGE = serum levels of soluble receptor for advanced glycation end products.

**Supplementary Figure 1**. GWAS quantile-quantile plots of observed versus expected -$\log_{10}$ ($p$-value) of kidney function (eGFRcr and eGFRcys) and sRAGE

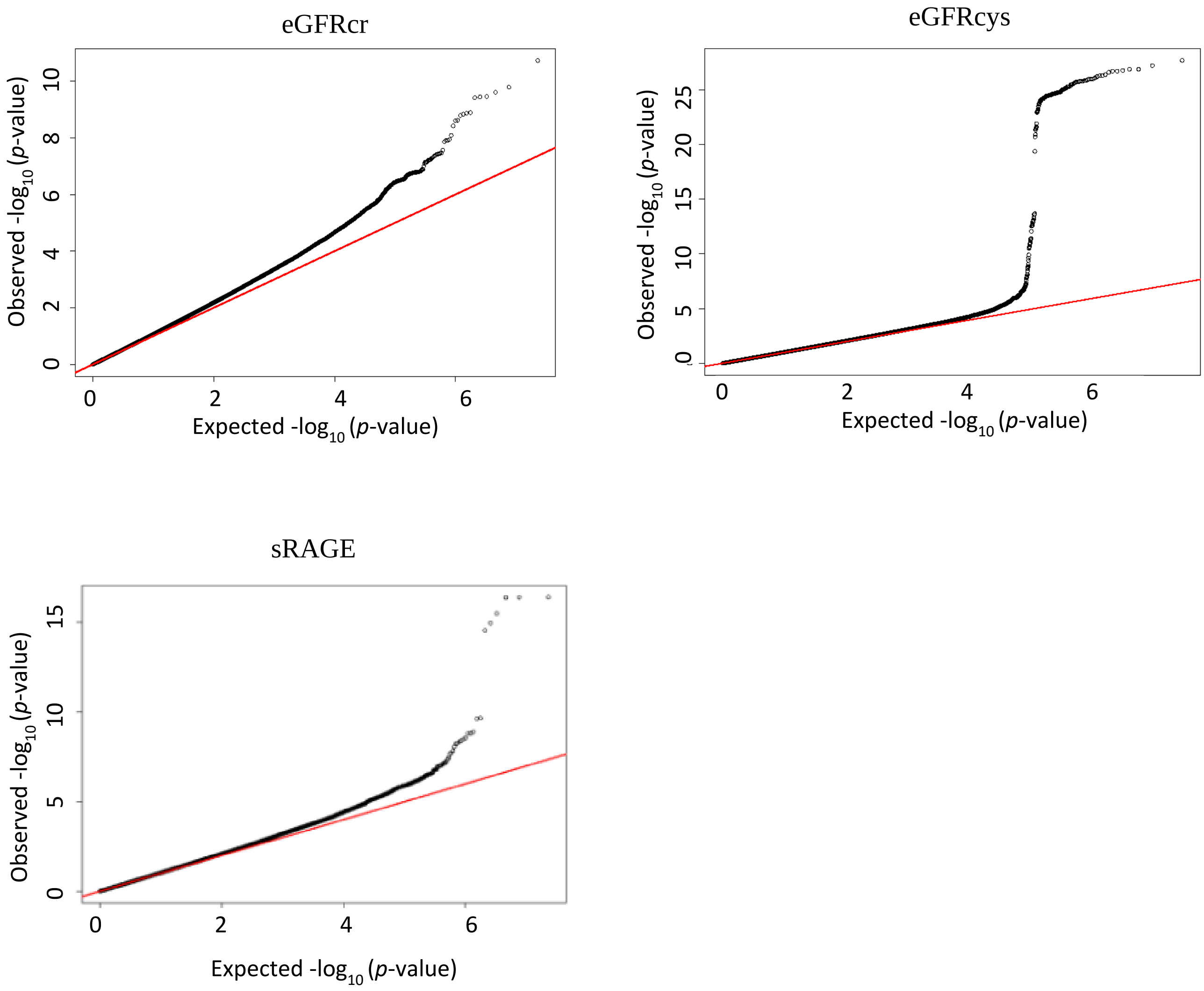

**Supplementary Figure 2.** Manhattan plots of univariate GWAS for kidney function (eGFRcr and eGFRcys) and sRAGE

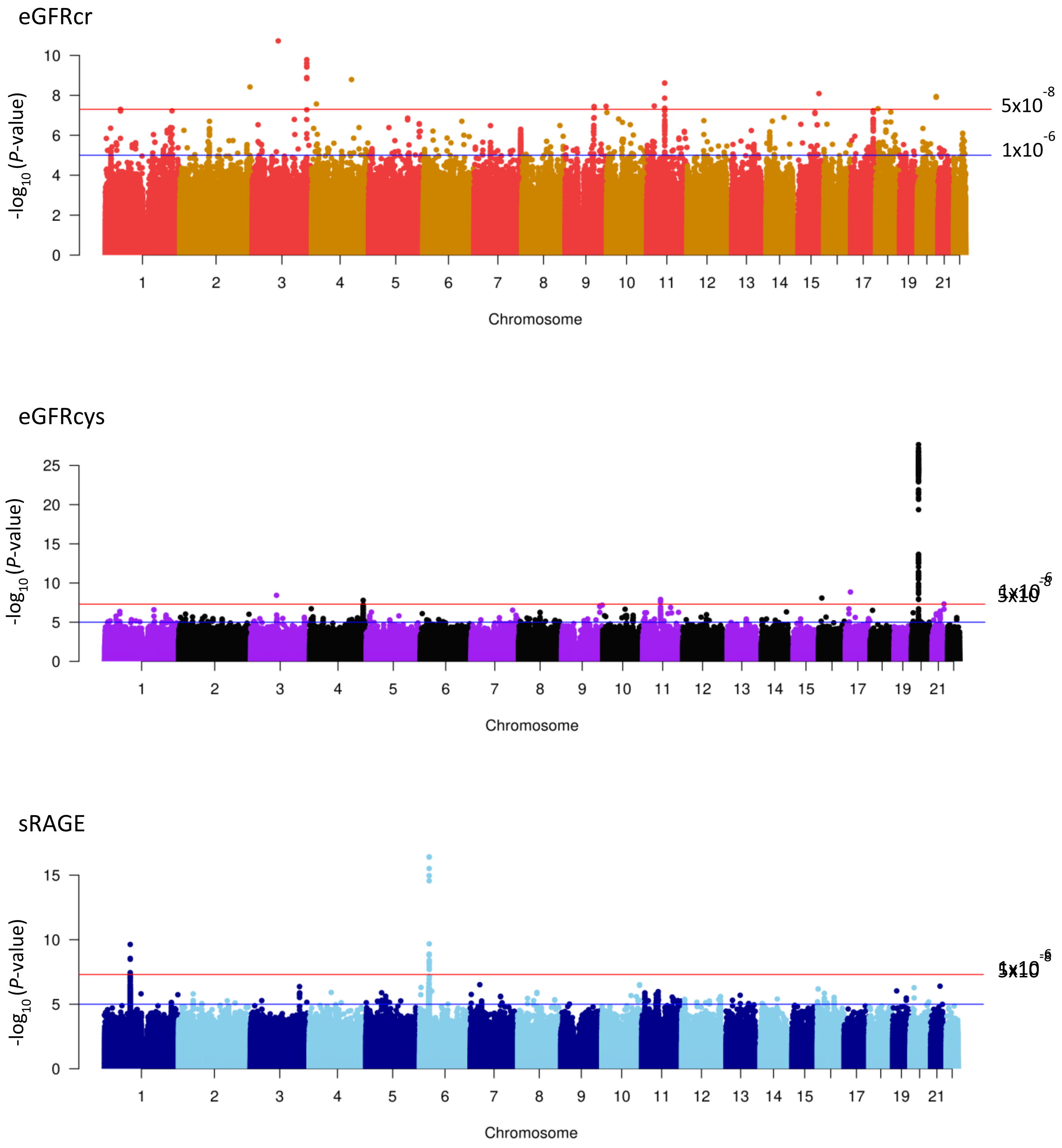

**Supplementary Figure 3.** Manhattan plot of CMA for kidney function (eGFRcr and GFRcys) and sRAGE

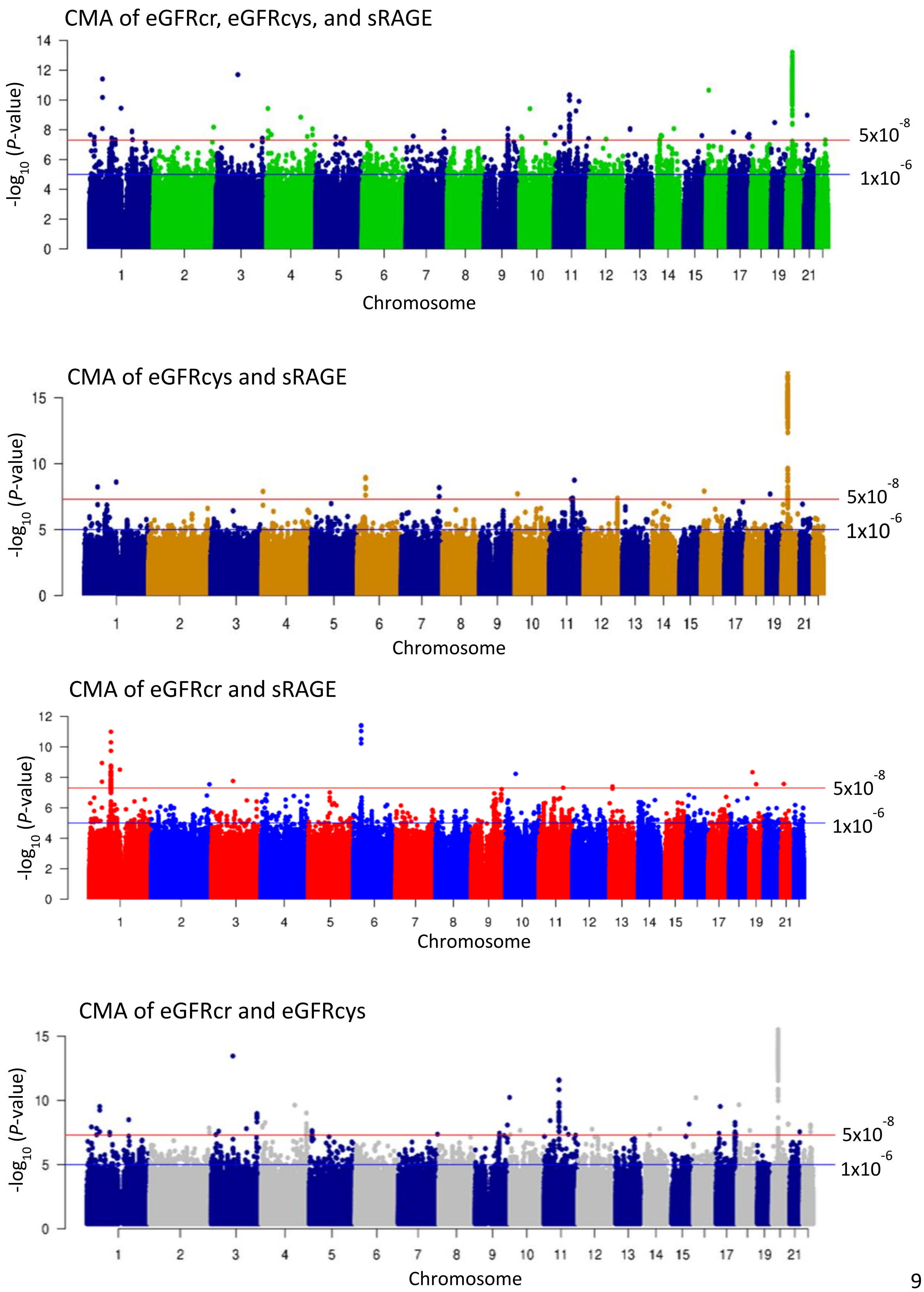

**Supplementary Figure 4.** Locuszoom plots of novel CMA GWAS

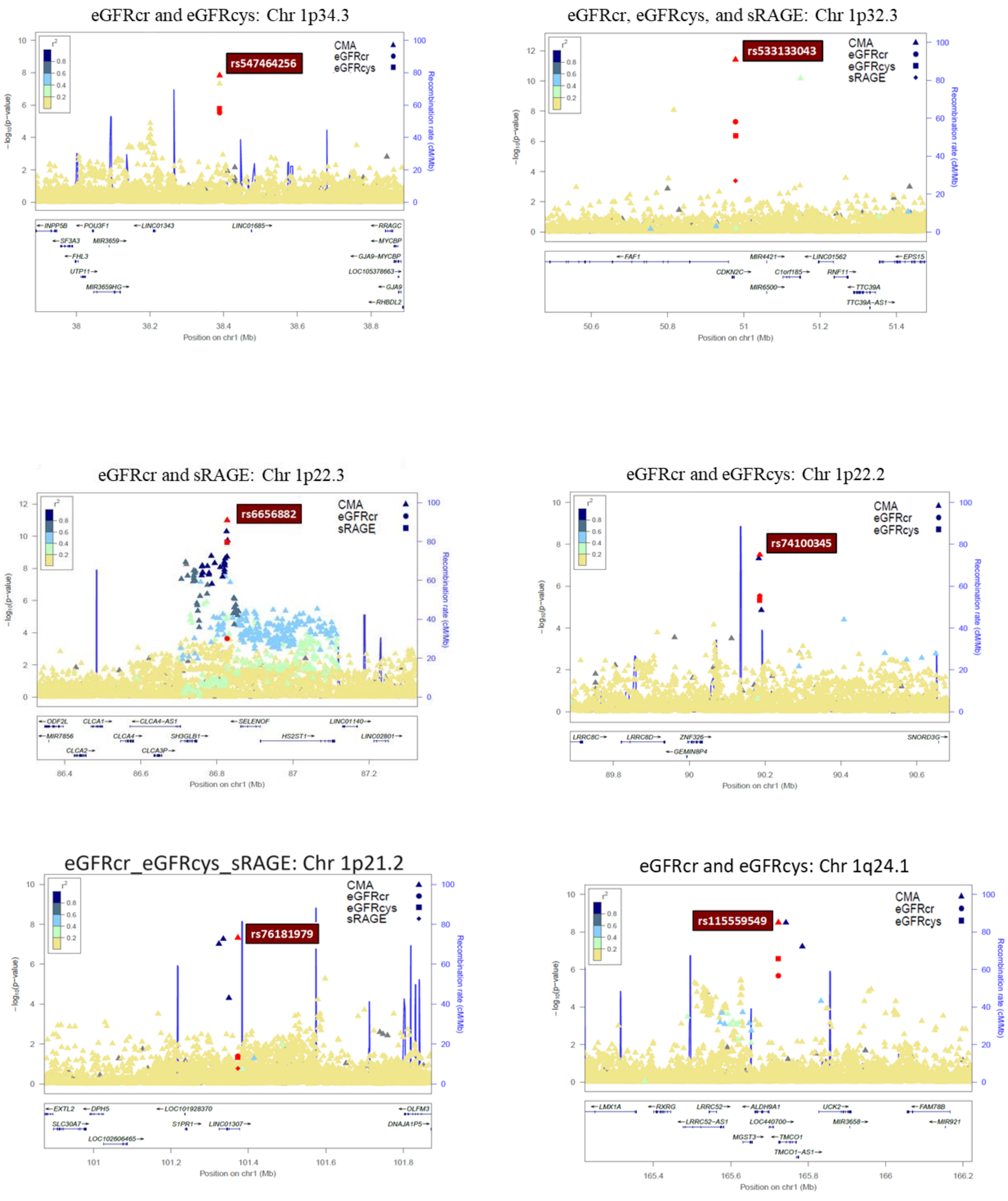

eGFRcr and eGFRcys: Chr 2q37.2

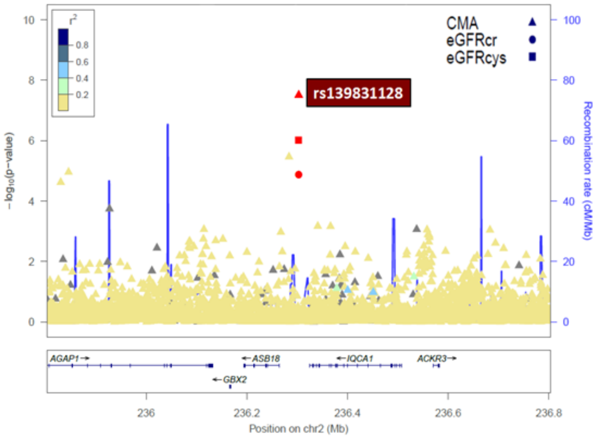


eGFRcr and eGFRcys: Chr 3p24.3

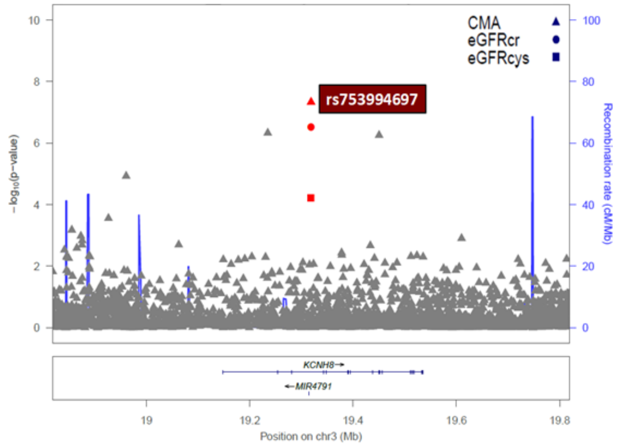


eGFRcr, eGFRcys: Chr 3p12.1

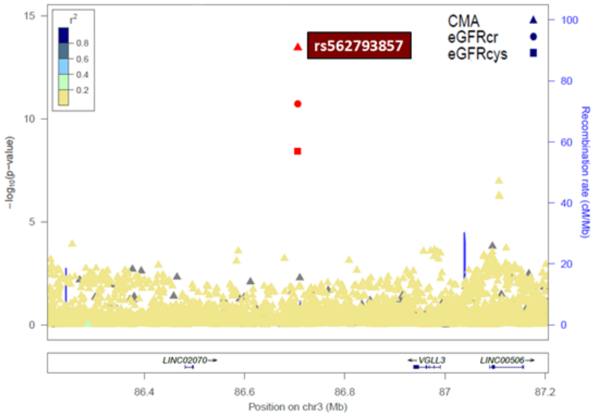


eGFRcr and eGFRcys: Chr 3q23

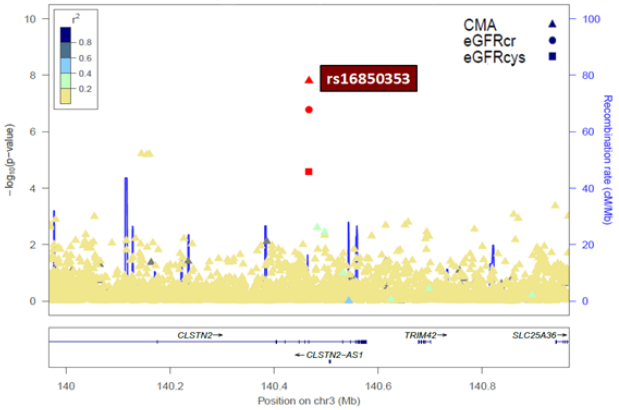


eGFRcr, eGFRcys, and sRAGE: Chr 4p16.2

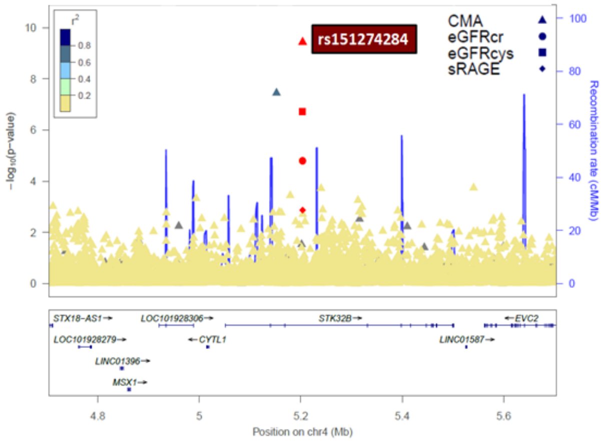


eGFRcr, eGFRcys, and sRAGE: Chr 4p15.31

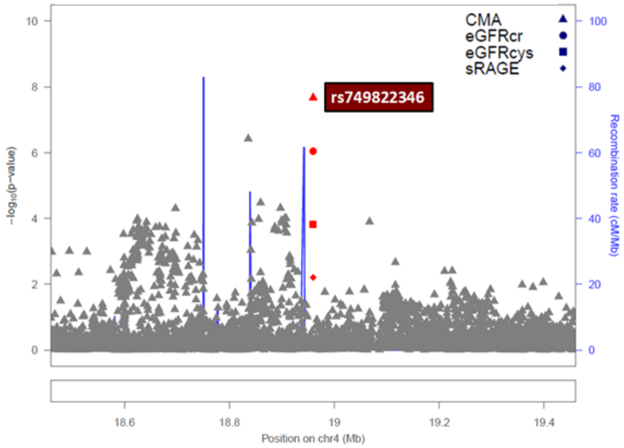

eGFRcr, eGFRcys: Chr 4q28.3

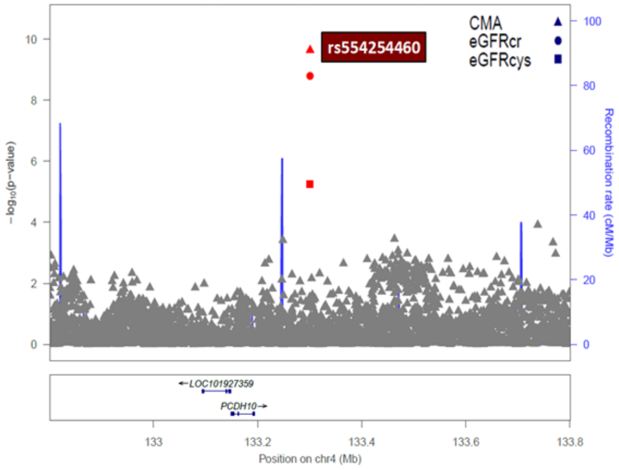


eGFRcr and eGFRcys: Chr 4q34.3

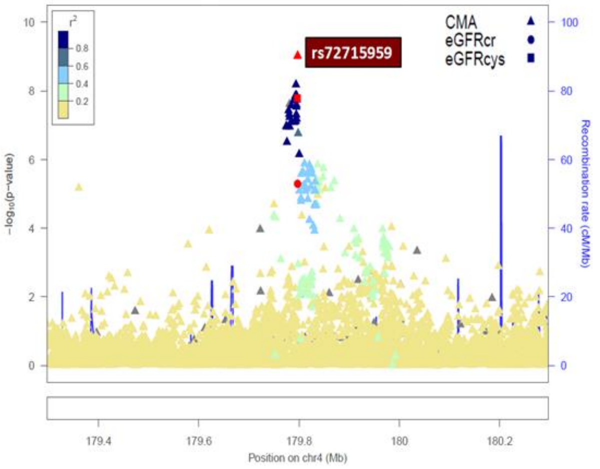


eGFRcr and eGFRcys: Chr 5p15.2

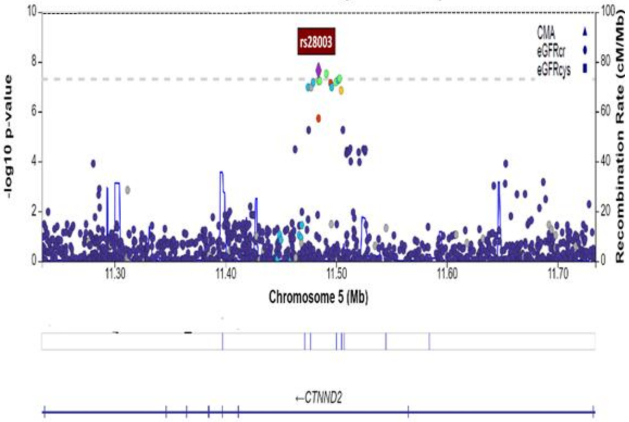


eGFRcr, eGFRcys, and sRAGE: Chr 5q14.1

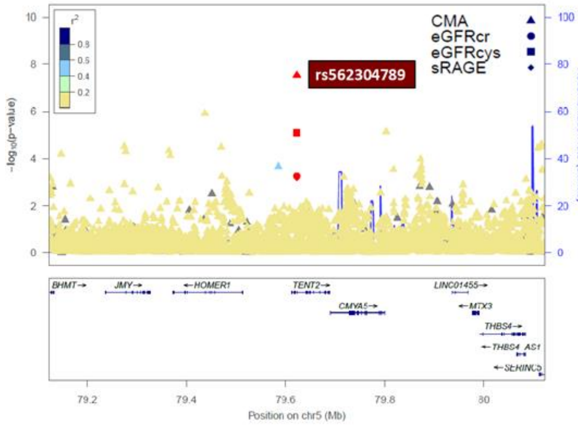


eGFRcr_eGFRcys_sRAGE: Chr 5q22.3

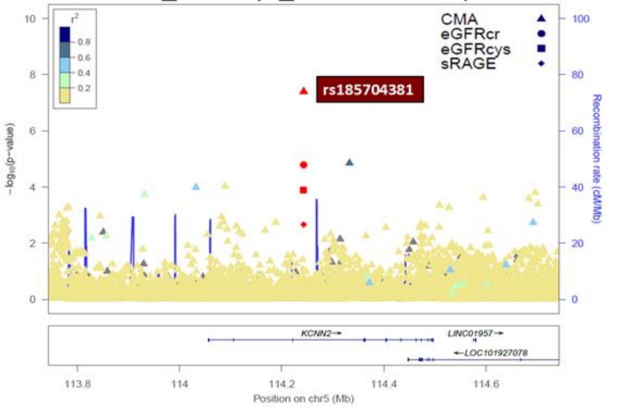


eGFRcr, eGFRcys, and sRAGE: Chr 7p15.1-p14.3

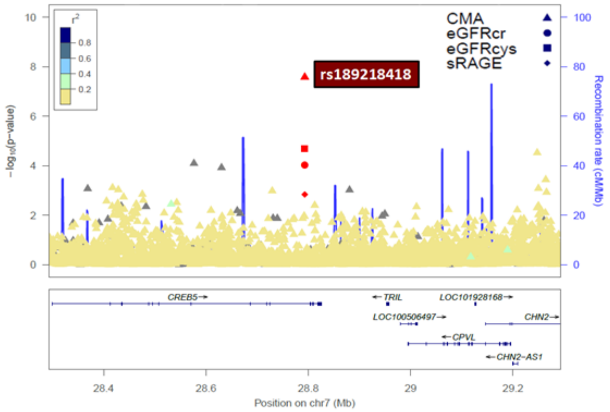

eGFRcys and sRAGE: Chr 7q35-q36.1

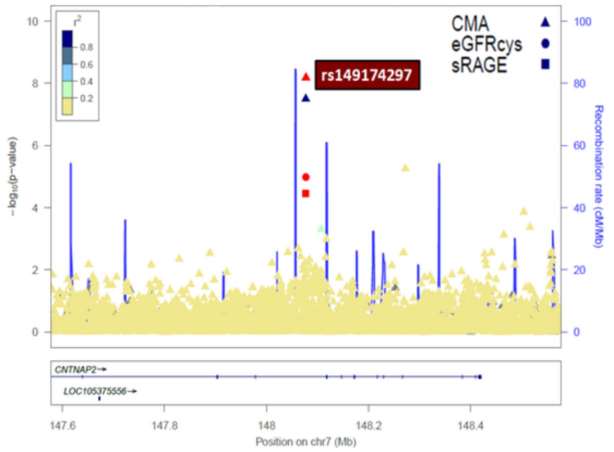


eGFRcr, eGFRcys, and sRAGE: Chr 9q22.31

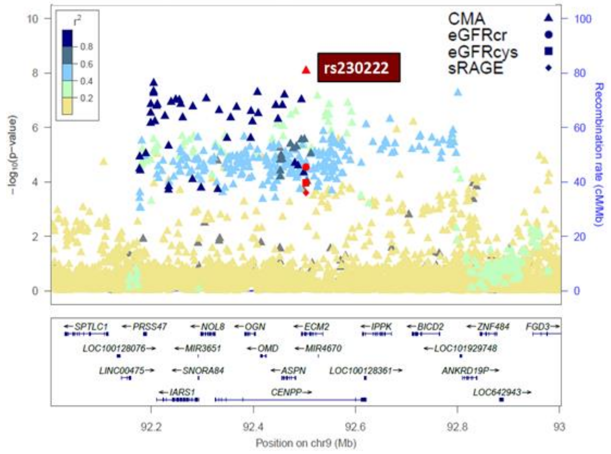


eGFRcr and eGFRcys: Chr 9q22.33

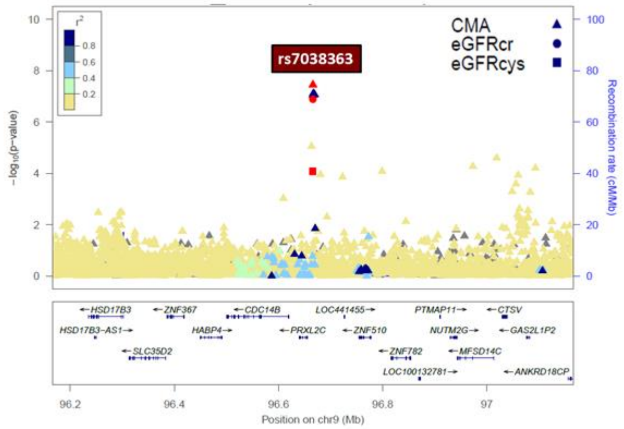


eGFRcr, eGFRcys, and sRAGE: Chr 10p14

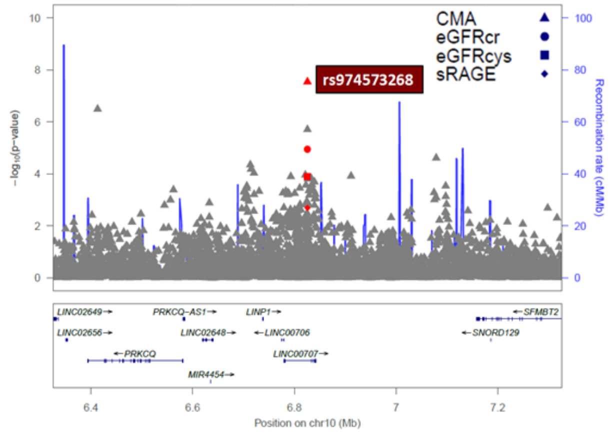


eGFRcr and sRAGE: Chr 10p14

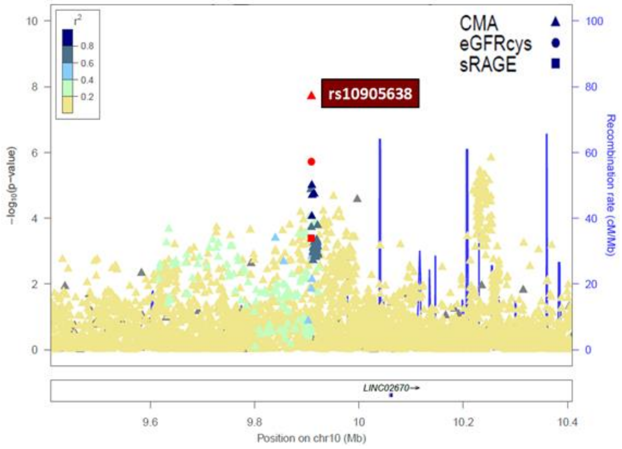


eGFRcr and eGFRcys: Chr 11p14.3-p14.2

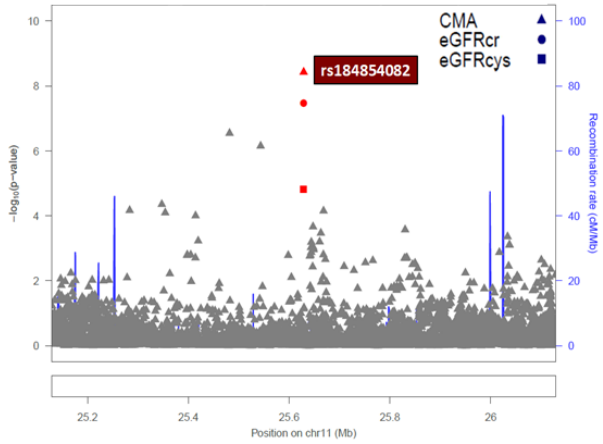

eGFRcr and eGFRcys: Chr 11q12.2

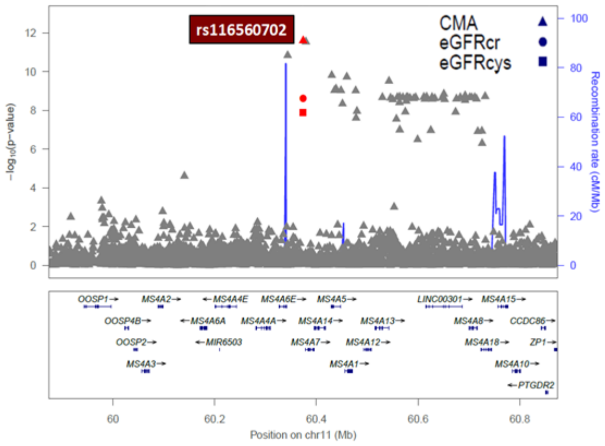


eGFRcr, eGFRcys, and sRAGE: Chr 11q14.1

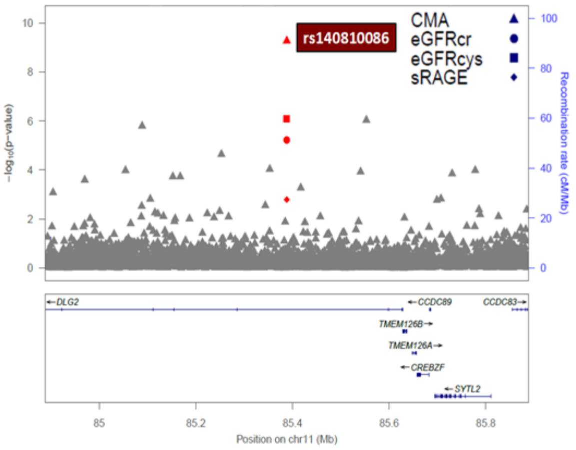


eGFRcr and sRAGE: Chr 11q14.3

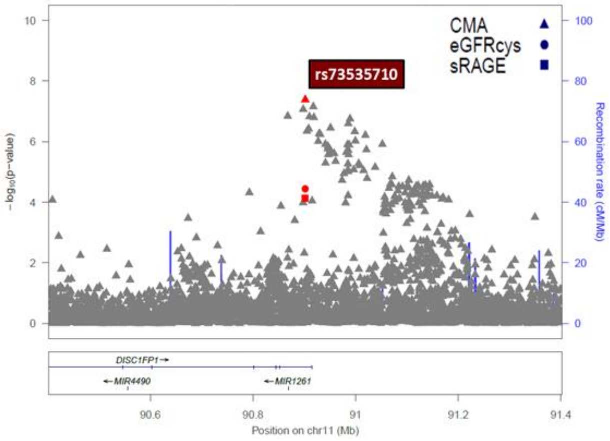


eGFRcr, eGFRcys, and sRAGE: Chr 11q22.1

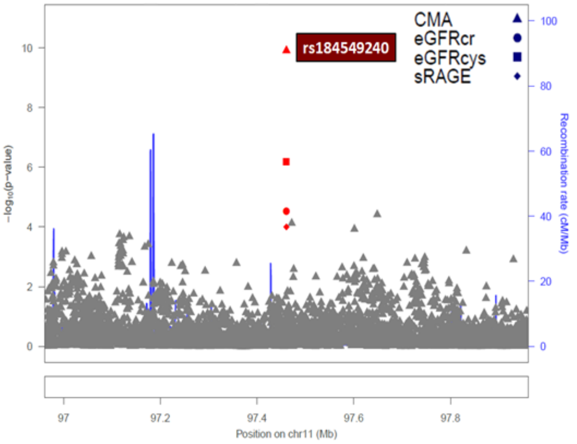


eGFRcr, eGFRcys, and sRAGE: Chr 11q25

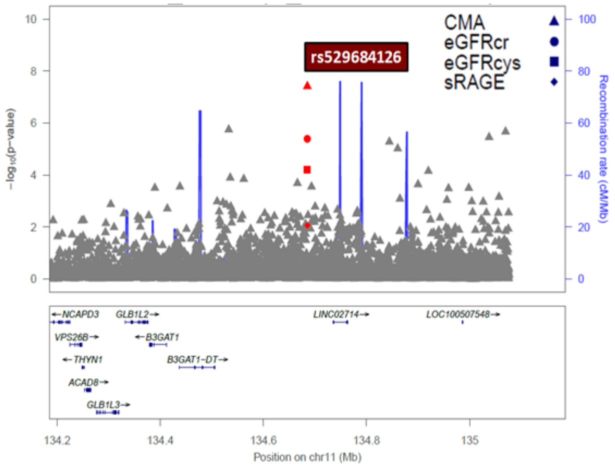


eGFRcys and sRAGE: Chr 12q24.33

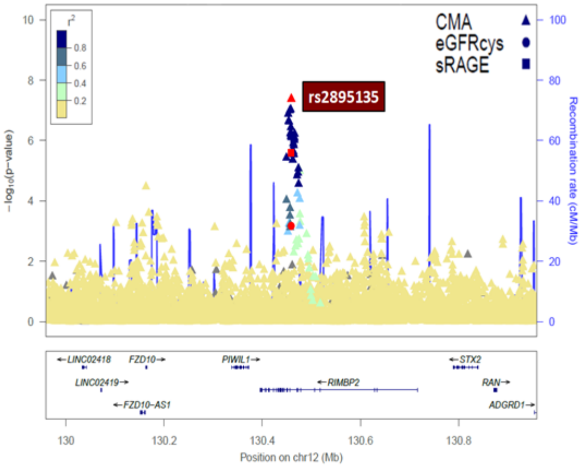

eGFRcr, eGFRcys, and sRAGE: Chr 14q12

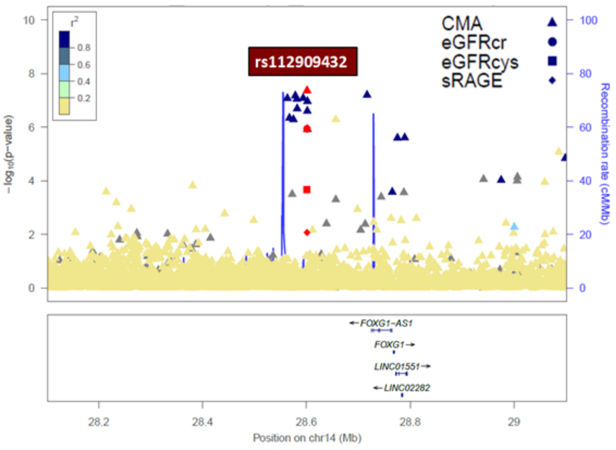


eGFRcr, eGFRcys, and sRAGE: Chr 14q13.1

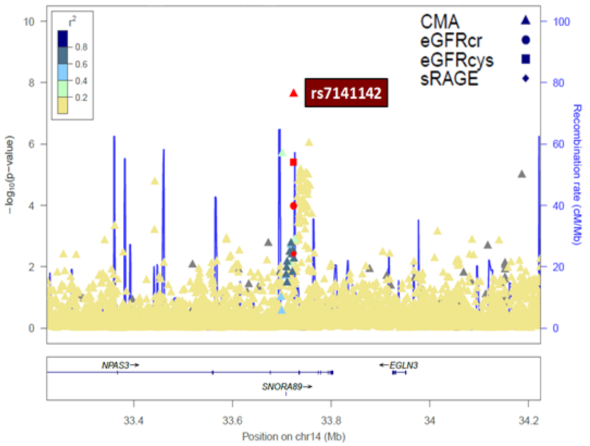


eGFRcr and eGFRcys: Chr 14q24.3

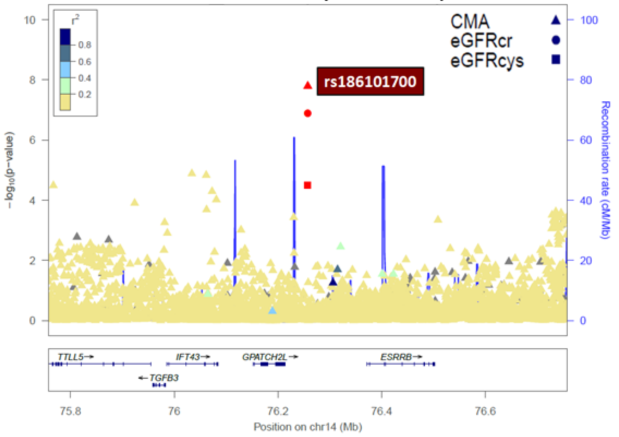


eGFRcr, eGFRcys, and sRAGE: Chr 14q31.3

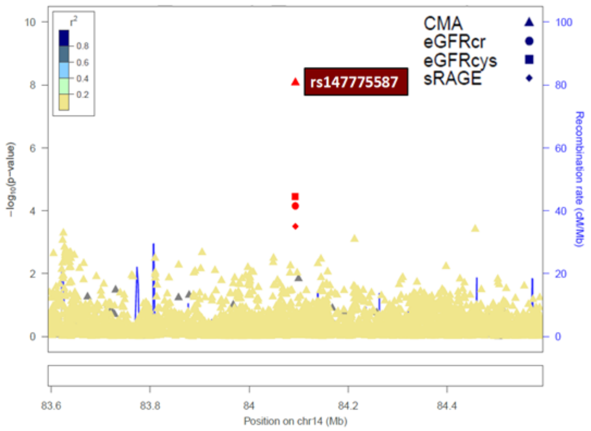


eGFRcr and eGFRcys: Chr 15q25.3

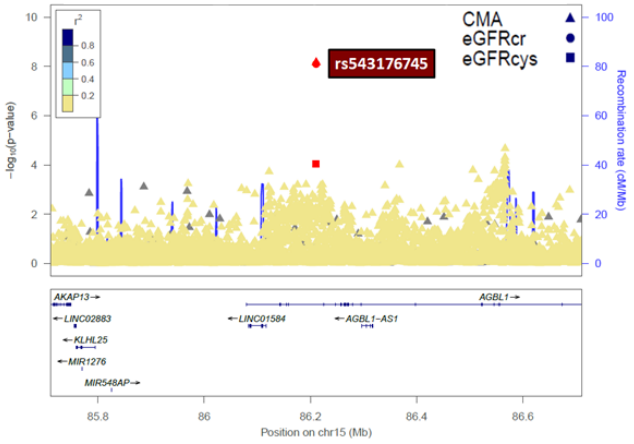


eGFRcr, eGFRcys, and sRAGE: Chr 17q24.3

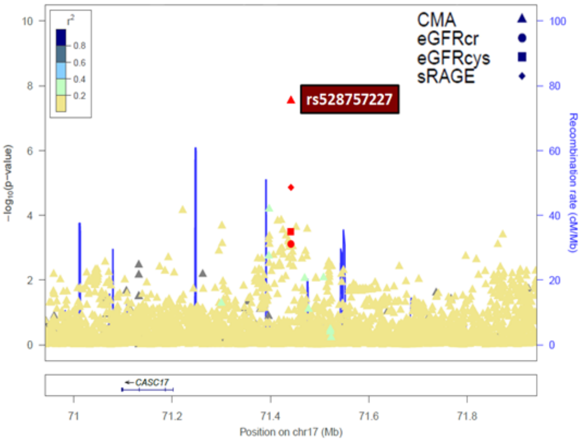

eGFRcr and eGFRcys: Chr 17q25.3

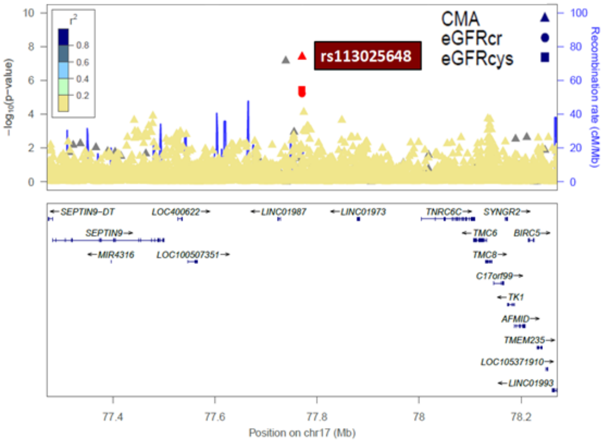


eGFRcr and eGFRcys: Chr 18p11.23

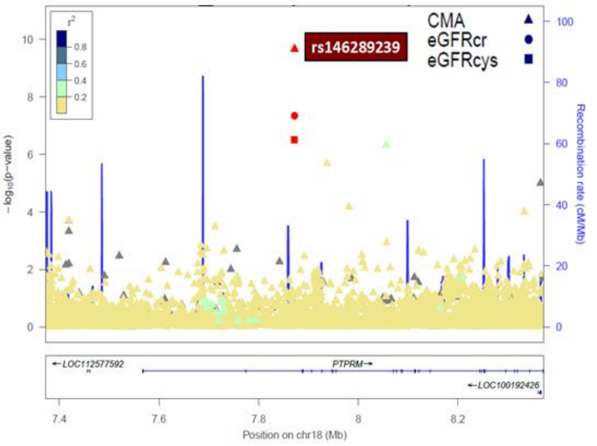


eGFRcr and eGFRcys: Chr 18q21.1-q21.2

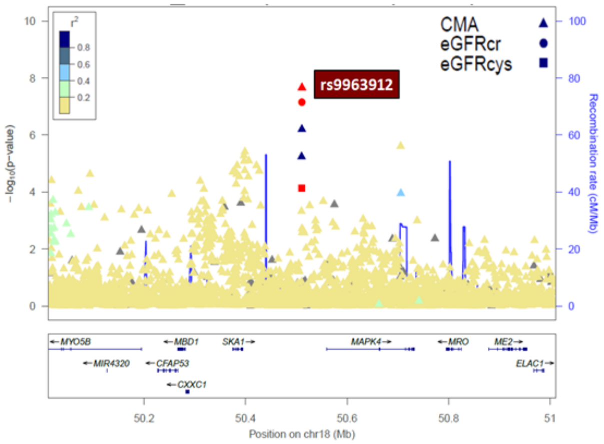


eGFRcr and sRAGE: Chr 19q12

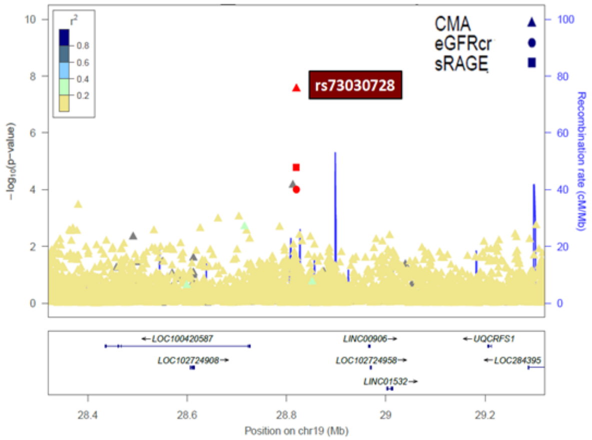


eGFRcr, eGFRcys, and sRAGE: Chr 21q21.1

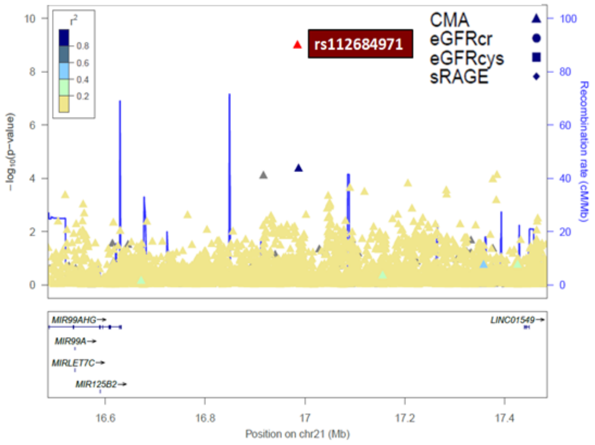


eGFRcr and eGFRcys: Chr 21q22.3

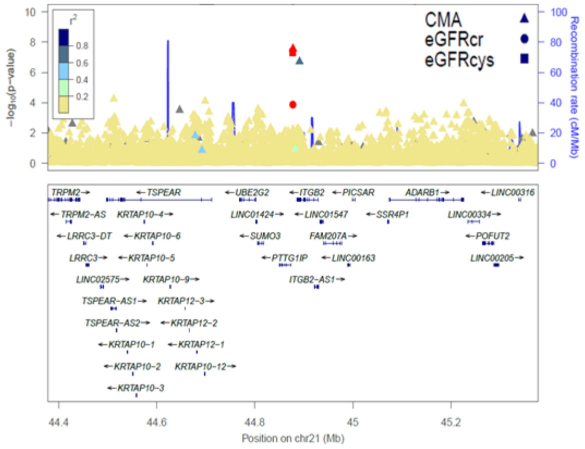

**Supplementary Figure 5.** TWAS quantile-quantile plots of observed versus expected $-\log_{10}$ (*p*-values)

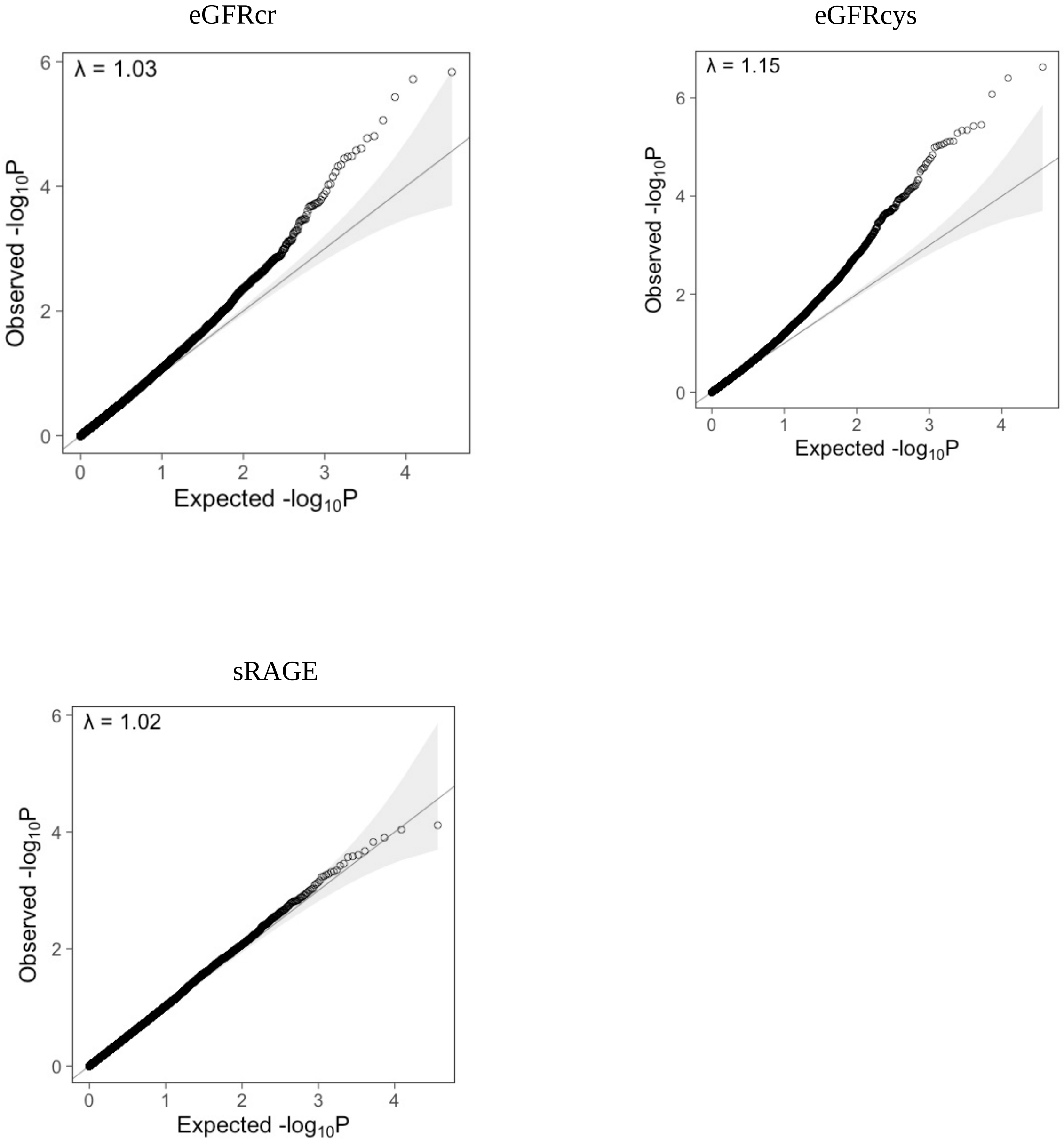
